# Optimal Deep Brain Stimulation Locations for Gilles de la Tourette Syndrome

**DOI:** 10.64898/2026.02.21.26346772

**Authors:** Ilkem Aysu Sahin, Konstantin Butenko, Kara A. Johnson, Helen Friedrich, Simón Oxenford, Ningfei Li, Patricia Zvarova, Barbara Hollunder, Nanditha Rajamani, Garance M. Meyer, Clemens Neudorfer, Maike Mustin, Lukas L. Goede, Anna Tietze, Wolf-Julian Neumann, Juan Carlos Baldermann, Till Dembek, Christina van der Linden, Anna C. von Olberg, Jens Kuhn, Daniel Huys, Pablo Andrade, Rabea Schmahl, Petra Heiden, Yulia Worbe, Nadya Pyatigorskaya, Carine Karachi, Marie-Laure Welter, Linda Ackermans, Anouk Y.J.M Smeets, Albert F.G. Leentjens, Chencheng Zhang, Bomin Sun, Jian-Guo Zhang, Fan-Gang Meng, Xinguang Yu, Xin Xu, Zhipei Ling, Domenico Servello, Alberto Bona, Mauro Porta, Alon Y. Mogilner, Michael H. Pourfar, Jill L. Ostrem, Thomas Foltynie, Patricia Limousin, Ludvic Zrinzo, Eileen M Joyce, Zinovia Kefalopoulou, Alida A. Postma, Luigi M. Romito, Matteo Vissani, Alberto Mazzoni, Osvaldo Vilela-Filho, Andres M. Lozano, Martin Parent, Abbas F. Sadikot, Kelly D. Foote, Christopher R. Butson, Veerle Visser-Vandewalle, Michael S. Okun, Andreas Horn

**Affiliations:** Einstein Center for Neurosciences Berlin, Charité – Universitätsmedizin Berlin, Berlin 10117, Germany; Institute for Network Stimulation, Department of Stereotactic and Functional Neurosurgery, University Hospital Cologne, Cologne, Germany; Movement Disorders and Neuromodulation Unit, Department of Neurology with Experimental Neurology, Charité – Universitätsmedizin Berlin, Corporate Member of Freie Universität Berlin and Humboldt-Universität zu Berlin, Berlin 10117, Germany; Center for Brain Circuit Therapeutics, Department of Neurology, Mass General Brigham, Harvard Medical School, Boston, MA, USA; Department of Neurology, Norman Fixel Institute for Neurological Diseases, University of Florida, Gainesville, FL, USA; Department of Neurology, University of Florida, Gainesville, FL, USA; Julius-Maximilian-University Wuerzburg, Germany; Northwestern Polytechnical University, China; Department of Neurosurgery, Massachusetts General Hospital, Harvard Medical School, Boston, MA, USA; Institute of Neuroradiology, Charité-Universitätsmedizin Berlin, Corporate Member of Freie Universität Berlin, Humboldt-Universität zu Berlin, and Berlin Institute of Health (BIH), Berlin, Germany; Department of Psychiatry and Psychotherapy, Medical Center - University of Freiburg, Faculty of Medicine, University of Freiburg, Freiburg, Germany; University of Cologne, Faculty of Medicine and University Hospital Cologne, Department of Neurology, Cologne, Germany; Department of Psychiatry and Psychotherapy, Faculty of Medicine and University Hospital Cologne, University of Cologne, Cologne, Germany; Alexianer Hospital Cologne, Alexianer Köln GmbH, Cologne, Germany; Department of Psychiatry and Psychotherapy III, LVR Klinik Bonn, Bonn 53111, Germany; Department of Stereotactic and Functional Neurosurgery, Faculty of Medicine and University Hospital Cologne, University of Cologne, Cologne, Germany; Sorbonne University, Inserm U1127, CNRS UMR7225, UM75, Paris Brain Institute, Movement Investigation and Therapeutics Team, 75013 Paris, France; Department of Clinical Neurophysiology, Hopital Saint Antoine, 75012 Paris, France; Sorbonne University, Paris Brain Institute - ICM, Inserm, CNRS, Department of Radiology, Hôpital de la Pitié Salpêtrière (DMU 6), AP-HP, 75013, Paris, France; Inserm 1127, Sorbonne Université, UPMC Univ Paris 06, UMRS 1127, CNRS, UMR 7225, Paris Brain Institute, Paris, France; Department of Neurophysiology, CHU Rouen, Rouen University, 76000 Rouen, France; Department of Neurosurgery, Maastricht University Medical Centre, 6229 HX Maastricht, The Netherlands; Department of Neurosurgery, Zuyderland Medical Center, Academic Neurosurgical Center Limburg, Heerlen, The Netherlands; Department of Psychiatry, Maastricht University Medical Centre, 6229 HX Maastricht, The Netherlands; Department of Neurosurgery, Rujin Hospital, Shanghai Jiao Tong University School of Medicine, Shanghai, China; Beijing Neurosurgical Institute, Capital Medical University, Beijing, China; Department of Neurosurgery, Chinese PLA General Hospital, Beijing, 100853, China; Department of Neurosurgery, Hainan Hospital of Chinese PLA General Hospital, Sanya, Hainan, 572000, China; Neurosurgical Department, IRCCS Istituto Ortopedico Galeazzi, Milan, Lombardia, Italy; Tourette’s Syndrome and Movement Disorders Center, IRCCS Istituto Ortopedico Galeazzi, Milan, Lombardia, Italy; Center for Neuromodulation, Departments of Neurology and Neurosurgery, New York University Medical Center, New York, New York, USA; Department of Neurology, University of California San Francisco, San Francisco, California, USA; Department of Clinical and Movement Neurosciences, UCL Queen Square Institute of Neurology, London, UK; National Hospital for Neurology and Neurosurgery, UCL Queen Square Institute of Neurology, London, UK; Department of Radiology and Nuclear Medicine, Maastricht University Medical Center, the Netherlands; Mental Health and Neuroscience Research Institute, Maastricht University, the Netherlands; Parkinson and Movement Disorders Unit, Fondazione IRCCS Istituto Neurologico Carlo Besta, Milan, Italy; The BioRobotics Institute, Scuola Superiore Sant’Anna, Pisa, Italy; Department of Excellence in Robotics and AI, Scuola Superiore Sant’Anna, Pisa, Italy; Division of Neurosurgery, Department of Surgery, Medical School, Federal University of Goiás, Goiânia, Brazil; Division of Neurosurgery, Department of Surgery, University of Toronto, Toronto, Ontario, Canada; Krembil Research Institute, Toronto, Ontario, Canada; CERVO Brain Research Centre, Quebec City, Quebec, Canada; Department of Neurology and Neurosurgery, Montreal Neurological Institute, McGill University Health Centre, McGill University, Montreal, QC, Canada; Department of Neurosurgery, Shands at University of Florida, University of Florida, Gainesville; Department of Neurosurgery, University of Florida, Gainesville, FL, USA; J Crayton Pruitt Family Department of Biomedical Engineering, University of Florida, Gainesville, FL, USA

## Abstract

**Background:** Deep brain stimulation has emerged as an effective investigational treatment for select cases of severe Gilles de la Tourette Syndrome. Defining the optimal stimulation sites within different targets and the specific tic improvement network across targets will be important to guide neuromodulation therapies.

**Methods:** This retrospective multi-center cohort study analyzed stimulation locations in patients who received bilateral deep brain stimulation for Gilles de la Tourette Syndrome across 12 centers world-wide. The brain targets included the thalamus (n=43), pallidum (n=56) and subthalamic nucleus (n=16). The median follow-up period was 6 months. Imaging data were processed using a dedicated pipeline and a recently introduced voxel-wise sweetspot mapping technique. Since tic response landscapes visually resembled streamline tract connections, we carried out extensive anatomical delineations of pallidothalamic and thalamostriatal fibers. This anatomical information was used to interpret sweetspot landscapes across the three target regions.

**Results:** Tic response maps revealed three tic-response peaks in both thalamus and pallidum. Based on thalamic and pallidal response maps, outcomes in the subthalamic DBS cohort, stimulated between the two other targets, could be explained (R=0.58, p=0.019). Across the three targets, response maps followed the anatomical course of three bundles. Namely, specific subregions of the ansa lenticularis, the fasciculus lenticularis, and projections from the posterior intralaminar thalamic nuclei to the lentiform nucleus. Stimulation overlaps with these bundles explained 19% of the variance in tic improvement across the three targets. Response maps could explain variance in an independent test cohort (n=8, R=0.70, p=0.026). Response maps were also calculated for obsessive compulsive behavior, which revealed similarities to the tic response sites in pallidum but clearly distinct results and generally less efficacy in the thalamus.

**Conclusion:** Our analyses identified tic response targets that followed the course of known structural projections interconnecting pallidum and thalamus.

## Introduction

Gilles de la Tourette Syndrome (GTS) is a neurodevelopmental disorder primarily characterized by sudden, involuntary motor and phonic tics. Tics are typically preceded by aversive premonitory urges. Frequently, GTS is accompanied by psychiatric comorbidities such as obsessive-compulsive behavior (OCB).^1^ In the last few decades, deep brain stimulation (DBS) has emerged as a treatment option for select patients with severe symptoms that may be insufficiently controlled using conservative therapy.^1–3^

DBS for GTS was introduced in 1999 by Visser-Vandewalle et al.^4^ and was applied to target intralaminar thalamic nuclei and the nucleus ventro-oralis internus, based on the successful results of lesion-based thalamotomies of the same region originally performed by Hassler and Dieckmann in the 1970s.^5,6^ The rationale included the idea of affecting the known projections from thalamus back to the striatum and the pallidum, as well as to the cortex. It was hypothesized that by disrupting these projections, the input to the striatum could be gated.

Since the original work in lesioning and later DBS, multiple targets have emerged within a network including the pallidum and intralaminar thalamic nuclei. Common targets included the intralaminar thalamic complex (Centromedian/Parafascicular, CM-Pf; Centromedian/Ventrooralis intermedius; CM-Voi), the anteromedial (amGPi) and posteroventral internal globus pallidum (pvGPi).^1^ The clinical outcomes from these surgeries have been promising. Meta-analytic evidence suggests a >50% reduction in tic severity in 238 out of 343 patients following DBS.^7^ Comorbid OCB symptoms also were found to improve, albeit generally less so with the CM-Pf target.^7^ Recent experimental targets such as the external globus pallidum (GPe),^8^ the subthalamic nucleus (STN)^9,10^ and the field of Forel H1^11^ have also shown promise.

Despite the overall positive clinical outlook, intra-target electrode implantation as well as clinical outcomes have been heterogenous across patients and centers.^7,12^ A consensus regarding the optimal target within thalamus or pallidum has not yet been reached.^12^ Moreover, it remains unknown where exactly to target, i.e. which specific subparts of each target site would lead to optimal stimulation response.

Besides the local target-specific effects, the network level organization underlying tic improvement across the numerous target regions also remains unresolved. Cortico-basal-ganglia-thalamo-cortical (CBGTC) circuits are implicated in the pathophysiology of tics and associated comorbidities in GTS.^13^ The observation that stimulation of different targets within this CBGTC circuit can reduce tics similarly^7^ suggests a potential shared therapeutic network. Electrophysiological evidence supports this notion. Simultaneous recordings from patients implanted with both bilateral CM and amGPi electrodes demonstrated that electrophysiological coupling *between these nuclei* is strongly associated with tic generation.^14,15^ From an anatomical perspective, the current GTS targets constitute interconnected nodes within the CBGTC circuitry.^16^ The STN, as well, is bordered by multiple white matter pathways, including pallidothalamic pathways traversing the fields of Forel en route from pallidum to thalamus.^16,17^ Targeting the STN has therefore been motivated, at least in part, by their proximity to critical white-matter pathways rather than by the nucleus alone.^9,11^ In parallel, shared pathways across DBS targets have been identified for other disorders such as obsessive-compulsive disorder^18^ and Parkinson’s disease.^19^ Whether tic improvement across GTS-DBS targets similarly results from the modulation of a common network, however, remains an open question.

From a functional perspective, GTS is a complex neuropsychiatric disorder involving symptoms that relate to both motor (i.e., the tics) and associative or limbic loops (i.e., the premonitory urge to tic and comorbidities).^1^ Consistent with this clinical heterogeneity, the GTS-DBS targets themselves are functionally segregated, encompassing motor, associative and limbic territories.^20^ This functional organization has direct clinical implications. For instance, more anterior lead placement within GPi may possibly impact more associative and limbic processing, while implantation in posterior regions seem to be associated with motor functions. Due to its complexity, GTS offers no *a priori* indication of which neural circuits are disrupted, nor whether symptoms stem from dysfunction within a *single* circuit or from imbalances across *multiple* circuits. Although optimal symptom control may require modulation of motor, associative and limbic components, the spatial extent of a single DBS electrode is inherently limited. Identification of critical network bottle necks (anatomical or functional convergence points) at which stimulation can exert broad effects across symptom spectrum could potentially impact treatment strategies.

Here, we leverage the largest-to-date retrospective GTS-DBS imaging cohort (*N*=115) including six DBS targets (amGPi, pvGPi, GPe, CM-Pf, CM-Voi, STN) across three major target regions: the thalamus, the pallidum and the STN. We address two complementary questions: (i) whether optimal stimulation sites for tics and OCB can be identified within individual DBS target regions and (ii) whether effective targets converge onto shared tract connections mediating tic improvement.

## Materials and methods

### Participants

Retrospective imaging data of 115 patients who had undergone bilateral DBS treatment for GTS, targeted in the thalamus (CM/Voi or CM/Pf; *n*=43), pallidum (amGPi, pvGPi, or GPe, *n*=56) or STN (*n*=16) across 12 centers world-wide were included (Supplementary methods, Supplementary Fig.1, Supplementary Table 1). The details of DBS eligibility criteria and surgical procedures can be found in each respective publication^8–10,12,21,22^. The study was approved by the institutional review board of Charité – Universitätsmedizin Berlin (IRB Number: EA2/186/18) and was conducted in accordance to the Declaration of Helsinki. Given the secondary use of deidentified research data, the study was exempted from obtaining informed consent.

**Figure 1:**
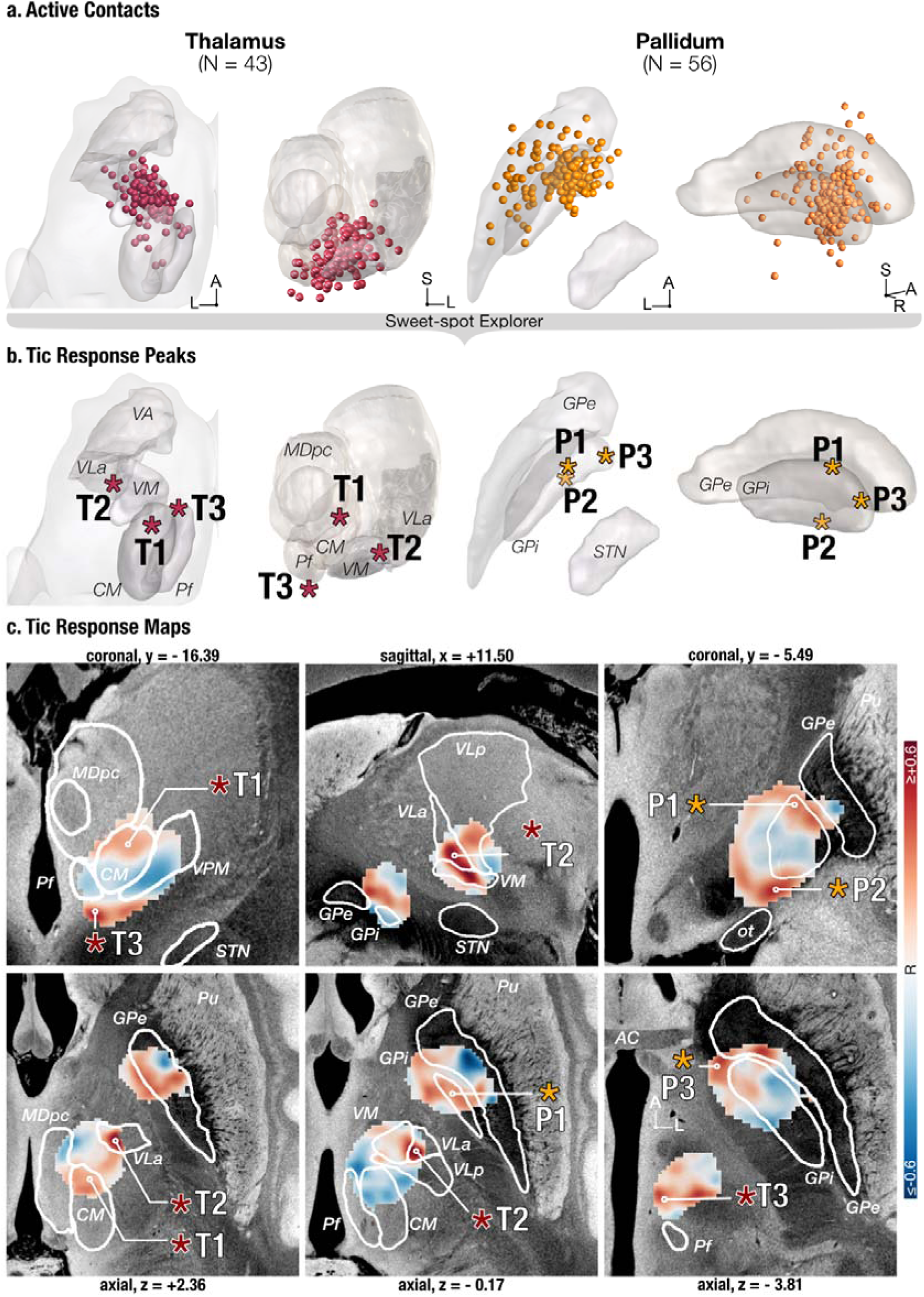
Local tic response maps of thalamus and pallidum. **a)** The active stimulation contact locations of thalamus (magenta) and pallidum (orange) cohorts, visualized in template space. 3D structures show GPe, GPi, STN and thalamic nuclei from FOCUS atlas^44^. Note the wide distribution of active contacts within each target nuclei. **b)** Voxel-wise correlations were calculated using the Sweet-spot Explorer as implemented in Lead-DBS v3.0.^26^ The main peaks of the tic response map were highlighted in 3D. **c)** Maps represent the local tic-response landscape, red indicating higher improvement with increased stimulation. Pink and orange stars represent thalamic (T1-3) and pallidal (P1-3) peaks, respectively. The color bar represents Pearson’s Correlation Coefficient. Images are visualized on top of a 7T image of an ex-vivo human brain.^46^ ***Abbreviations.*** *Globus Pallidus Externa, GPe, Globus pallidus interna, GPi, Ventrolateralis Anterior, VLa, Ventromedial Nucleus, VM, Ventral Posteromedial Nucleus, VPM, Centromedian, CM, Parafasciculus, Pf, Mediodorsal pars parvicellularis, MDpc, Subthalamic Nucleus, STN*.

DBS targets were CM/Pf/Voi regions of thalamus (*n*=43), amGPi (*n*=42), GPe (*n*=7), pvGPi (*n*=5), GPi/GPe border (*n*=2) and STN (*n*=16). Electrical fields (E-fields) of patients that are targeted at amGPi, pvGPi and GPe were grouped under “Pallidum”, due to the considerable overlap of E-fields across nuclei, similar to reports from Johnson et al.^23^ All thalamic targets (CM/Voi or CM/Pf) were also gathered under “Thalamus”, considering the expanse of the E-fields showing continuous coverage on a group level across multiple thalamic nuclei and the variability in targeting and thalamic nomenclature, as previously also noted by Wehmeyer *et al*.^7^ As a result, we had three anatomical cohorts in our analysis, thalamus (n=43, 13 females), pallidum (n=56, 15 females) and STN (n=16, 3 females).

Follow up time-points were harmonized at six months or the nearest follow-up date. Tic outcome was defined as the percent change of the available tic score, either Yale Global Tic Severity Scale (YGTSS, n=90)^24^ or Total Tic Severity Scale (TTSS, n=25), which involved the motor and phonic tic scores of the larger YGTSS. A subgroup of patients (n=38, 13 females; n=16 in thalamus, 5 females; n=22 in pallidum, 8 females) also had preoperative and postoperative Yale-Brown Obsessive Compulsive Scale (Y-BOCS)^25^ measurements available. Change in obsessive compulsive comorbid symptoms was defined as the percent change in Y-BOCS.

### Image Processing

Patient scans were preprocessed following the established default image analysis pipeline available in Lead-DBS v3.0 (https://www.lead-dbs.org/, ^26^). For 105 patients, the individual anatomy was based on preoperative MRI images. The electrode localizations for these patients were informed by postoperative CT (*n*=71) or MRI (*n*=34) images. In cases for which only postoperative MRI images were available (*n*=10), both the anatomy and the electrode localizations were informed by postoperative MRI scans. Preoperative and postoperative scans were coregistered using Advanced Neuroimaging Tools (ANTs, http://stnava.github.io/ANTs/)^27,28^ for postoperative CT scans and SPM12 (http://fil.ion.ucl.ac.uk/spm/)^29^ for MRI scans. Next, all images were normalized to standard template space (ICBM 2009b nonlinear asymmetric template)^30^ with either ANTs^27,28^ or EasyReg^31,32^ using default parameters implemented in Lead-DBS and refined using WarpDrive^33^, if necessary. For the 10 cases with only postoperative MRI scans, postoperative images were normalized to the standard space, and the rest of the pipeline was carried out in the same way with the rest of the cohort. For all patients, postoperative images were corrected for brain shift using a coarse mask, to account for potential tissue displacement due to pneumocephalus.

### Electrode Localization and E-field Modelling

After preprocessing, electrode trajectories were reconstructed in native patient space using PaCER^34^ or TRAC/Core^35^ algorithms available under Lead-DBS and manually refined if necessary. Tissue segmentation masks were created with SynthSeg^36^ and visually inspected for each patient. Then, E-fields were modeled isotropically in native space, using the pulse-width adaptive approach, as implemented in OSS-DBS^37^. E-field modelling was based on the respective stimulation parameter corresponding to the follow-up timepoint. The following conductivity values were used: gray matter, 0.115 S/m; white matter, 0.070 S/m; cerebrospinal fluid, 2.000 S/m. To allow for group-level analyses, the transformation matrices generated in the previous steps were used to transform E-fields of each patient from native space to standard template space. The rest of our analyses were based on E-fields. Volumes of tissue activated (VTAs) were used only for visualization.

### Voxel-wise Correlations: DBS Response Maps

The main goal of our study was to investigate the optimal stimulation sites for tics and OCB improvement. To do so, we applied DBS sweetspot mapping following the approach of ^38,39^. DBS response (sweetspot) maps were generated to explore correlations between applied electricity (E-field magnitude) and tic/OCB improvement on a group level. For this analysis, E-fields were mirrored across hemispheres and compiled together in template space. Voxels that were stimulated by a minimum of 0.2V/mm^40^ by at least 6 E-fields were included in the analysis. Critically, this does not mean that correlations were carried out across a minimum of six data points only, since numerous additional E-fields would stimulate any given voxel with less than 0.2V/mm. The value of 0.2V/mm has often been used as a heuristic ‘activation threshold’ in binarized volume of tissue activated analyses^38–40^ and was hence adopted here, as well. The resulting map highlighted areas in which stronger stimulation was associated with better (sweetspot) or worse (sourspot) clinical outcomes. The pipeline for these analyses is openly available in the form of the Sweet Spot Explorer tool as implemented in Lead DBS v3.0^26,38^. Once maps were defined, they could be used to infer the improvement of individual patients (either in-sample or out-of-sample from independent cohorts) as done previously^38,39^. To do so, E-fields of individual patients were overlayed with the response map. E-field intensities at the overlapping voxels were spatially correlated using percentage bend correlation (which is a robust form of correlation less prone to overfitting), yielding a “sweet-spot score” (expressed in form of a correlation coefficient) for each patient. Sweet-spot scores for each patient were then correlated with the respective empirical tic improvements.

### Candidate Tic Response Tracts

Since the resulting tic response map followed anatomical fiber tract formations that traversed between pallidum and thalamus (Video S1), we wanted to test empirically which structural connections contributed most to DBS response. To do so, we created a subcortical white matter atlas that included potential tract connections in the area that are known from the literature (see extensive supplementary material, Supplementary Figs. 3-9). Comparing sweetspot maps with these anatomical bundles and discussions with expert anatomists (M.P., A.S.) led us to assign the tic response peaks into sub-bundles of three anatomical pathways. Each candidate pathway was modelled manually in the form of curves in 3D Slicer, following the tract-like voxel clusters, while also adhering to the anatomically defined trajectories. 200 uniformly distributed streamlines were generated around the curve models, using CurveToBundle (Supplementary Fig.2, Supplementary Methods). This step resulted in three tic response tracts, based on voxel-wise correlations in the previous step and anatomical evidence.

**Figure 2:**
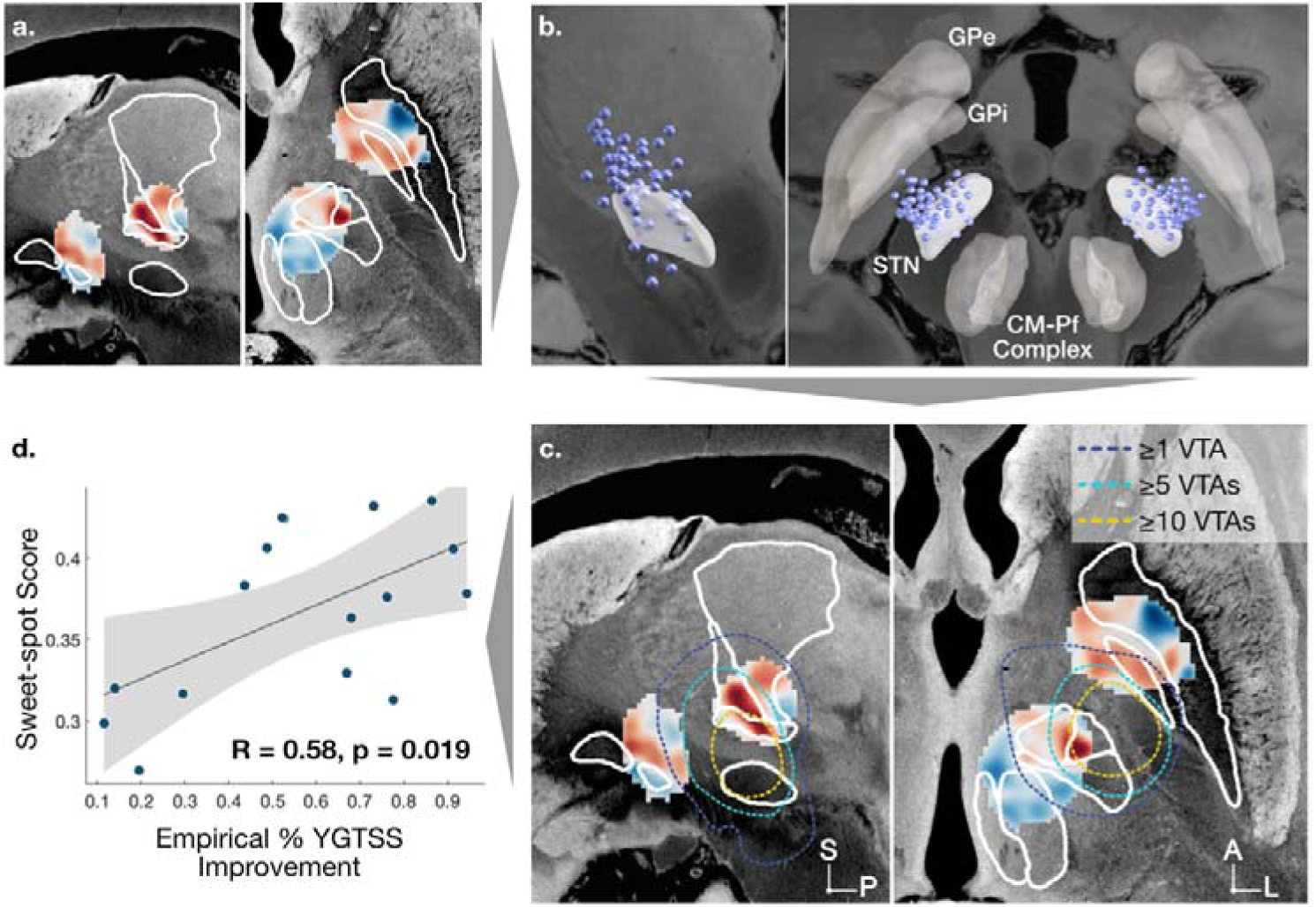
Thalamus and pallidum maps explain variance in STN cohort. **a)** The sweetspot map of thalamus and pallidum patients were spatially correlated to E-fields of STN patients (n=16). **b)** Active contacts of STN cohort. **c)** N-maps of stimulation volumes for STN patients are shown with dashed relief contour lines overlaid on the combined response landscapes of thalamic and pallidal cohorts. This shows that many patients, originally stimulated with STN as target, had significant overlap of their stimulation fields with sweetspot regions defined in the pallidal or thalamic cohorts. **d)** Spatial correlation between STN-DBS E-fields and the pallidothalamic tic response map led to one coefficient per patient, which was used as independent variable to explain variance in clinical outcomes of the STN cohort (dependent variable). ***Abbreviations.*** *Globus Pallidus Externa, GPe, Globus pallidus interna, GPi, Ventrolateralis Anterior, VLa, Ventromedial Nucleus, VM, Ventral Posteromedial Nucleus, VPM, Centromedian, CM, Parafasciculus, Pf, Mediodorsal pars parvicellularis, MDpc, Subthalamic Nucleus, STN*.

To quantify how much variance in clinical outcomes these tracts could account for, we first calculated E-field overlaps with the tracts and correlated these metrics to clinical outcomes. When further correlating the electrical field magnitude imposed on each streamline by each E-field with the respecting clinical outcome, we could assign a single R and p-value to each tract (following the ‘fiber filtering’ concept introduced earlier^41^). These values were used to further refine the candidate tract models by significance (*p*<0.01). Critically this represented a circular design, since the fibers had already been delineated along the sweetspot data that were informed by all patients. We nonetheless carried out the analysis to define a single tract model that could explain a maximum amount of variance across the three target sites.

### External Validation

To validate the sweetspot model generated from the main cohort (*N*=115), we identified an independent subset of 8 GTS patients retrospectively treated with DBS (CM/Voi, *n*=8) and followed at our local center (University Hospital Cologne) for whom complete clinical and imaging data were available. These patients were not part of the previous publications and have not been included in prior analyses (Supplementary Fig. 1). Detailed clinical and demographic information are presented in Supplementary Table 1. The preoperative MRI images and the postoperative CT scans were processed and E-fields were calculated with the exact pipeline described above.

### Data availability

The tic and OCB response maps are made openly available as an atlas within Lead-DBS (www.lead-dbs.org), which allows direct application in prospective or retrospective research studies that could validate/falsify the results. Lead-DBS also includes a streamline atlas that integrates decades of evidence from tracing studies and postmortem segmentations, assembled as tract models in standard template space, openly available to the scientific community. It may offer a reference for future DBS research across disorders such as epilepsy^42^ and consciousness modulation^43^. The same atlas streamlines were also contributed to the FOCUS atlas^44^, which is a comprehensive atlas of the subcortex curated by our institute.

### Code availability

CurveToBundle provides a tool for anatomically grounded pathway reconstruction and is openly available under the SlicerNetstim extension (https://github.com/netstim/SlicerNetstim.git) in 3D Slicer (https://www.slicer.org/).^45^

## Results

### Demographics and Clinical Scores

This study included a retrospective multi-center cohort of 115 patients who received bilateral DBS electrode implantation for GTS, targeted in the thalamus (*n*=43; CM/Voi, CM/Pf), pallidum (*n*=56; amGPi, pvGPi, GPe) or STN (*n*=16), establishing the largest and most comprehensive imaging dataset for GTS-DBS to date. Mean age at surgery was 29.5 ± 10.4 years. Average tic improvement after DBS implantation was 45.1% ± 26.6 at a median follow-up of 6 months [IQR: 6-8]. Detailed demographic and clinical information can be found in Supplementary Table 1.

In patients for which Y-BOCS scores were available (*n*=38), average age at DBS implantation was 30 ± 9.1 years. Mean Y-BOCS improvement was 17.4% ± 35.3 at a median of 6 months [IQR: 6-10]. The pallidal cohort improved significantly stronger compared to the thalamic cohort (Welch’s t(23.4) = 2.64, p=0.015); in fact, only three of the 16 thalamic patients had greater than 15% Y-BOCS improvement (while in the pallidum, 15/22 improved >15%). Y-BOCS improvement correlated significantly with tic improvement in the pallidal, but not in the thalamus cohort. (Supplementary Fig. 10)

### DBS Response Maps

Using DBS sweetspot mapping^38^, we analyzed the local DBS effects across targets, aiming to investigate the tic and OCB response maps within each target. The positive voxel clusters of thalamus (*n*=43) and pallidum (*n*=56) yielded three main response peaks in both thalamus and pallidum (T1-T3 and P1-P3; Fig. 1). For thalamus, the dorsal-most peak, T1, was located on the border of rostrodorsal CM and parvocellular component of mediodorsal thalamic nucleus (MDpc). T2 covered ventromedial (VM) and ventrolateralis anterior thalamic nuclei (VLa). Lastly, T3, the ventralmost peak, was located ventrally to Pf, in between Pf and nucleus ruber, at the ventral exit zone of intralaminar efferent connections. For pallidum, P1 covered the dorsalmost region of mid-thirds of GPi. P2 was observed in ventral GPi, overlapping with the caudalmost exit point of histologically observable ansa lenticularis^44^. P3 was located on amGPi at the level of the anterior commissure. (Fig. 1c) Additionally, positive voxel clusters resembling streaks between GPe and GPi were observed on axial slices, located laterally adjacent to P3 in anteromedial GPe and at mid-levels bordering P1. (Fig. 1c, Slices z = -3.81 and z = -0.17)

The OCB response peak in the pallidum (*n*=22) resided in the dorsal GPi, partly overlapping with the dorsal tic improvement peak, P1, and extended slightly more anteromedially (Supplementary Fig.11). In the thalamus (*n*=16), however, the OCB peak did not overlap with any of the major tic response peaks but instead resided in the intermedullary lamina between Centralis Medianus (CeM), CM and VM thalamic nuclei. Critically, as mentioned before, this peak in thalamus was driven by only the 3 patients who showed above >15% improvement in this cohort and should hence not be overinterpreted (Supplementary Fig. 12). These data may however confirm the general trend that sites modulating distinct symptoms may be more clustered in the basal ganglia vs. more segregated in the thalamus^19,47^.

Since the STN target is situated in between the two more established targets, it embodied an ideal first test whether fibers of passage (traversing between pallidum and thalamus), instead of a specific effect mediated by the STN itself, would account for tic relief after DBS. For this reason, STN cases had been left out in the initial sweetspot models (Fig. 1). STN-DBS E-fields were spatially correlated with the tic response map calculated from the thalamus and pallidum cohorts, leading to a correlation coefficient for each STN electrode, averaged across two hemispheres. Higher coefficients were calculated in cases where an STN E-field gradient followed the spatial pattern of the pallidothalamic tic response map, while E-fields that did not follow the spatial gradient of cohort-derived peaks yielded lower or negative coefficients. These coefficients reflected how well stimulation volumes of the STN cohorts matched the pallidothalamic tic response maps, and significantly correlated with clinically measured tic improvements of the STN patients (*R*=0.58, *p*=0.02; *n*=16; Fig. 2). In brief, the model derived from pallidum and thalamus cohorts was able to account for clinical outcomes in the entirely independent STN cohort.

After this analysis, the tic response map of the STN cohort (*n*=16) was also calculated. The tic response peak was located dorsally to the anterior STN, mainly outside the nucleus and extending into zona incerta (Supplementary Fig. 13).

### DBS Response Pathways

Fig. 3A shows the tic response landscape across all patients (*N*=115), which revealed that optimal stimulation sites followed known anatomical tract connections between pallidum and thalamus (fig. 3; also see video S1). In direct comparison with our independently created anatomical *a priori* tracts (see extensive supplement) and based on intensive discussions with expert anatomists (see the acknowledgement section), the peak clusters aligned with specific sub-components of three pathways. Namely, components of ansa lenticularis, fasciculus lenticularis and the outflow collaterals projecting from intralaminary thalamic nuclei—most likely Pf and/or medial CM—into GPi/GPe along their way to ventral putamen were identified (Fig. 3). At the dorsal regions of STN, ansa lenticularis and fasciculus lenticularis components of the candidate tic response bundles were both situated more posteriorly than the ‘canonical’ motor textbook definitions of the two tracts, mainly converging in extensions of Forel’s fields that resided close to zona incerta before entering associative thalamic areas (VM/VLa; for a direct comparison, see Supplementary Fig. 14).

**Figure 3:**
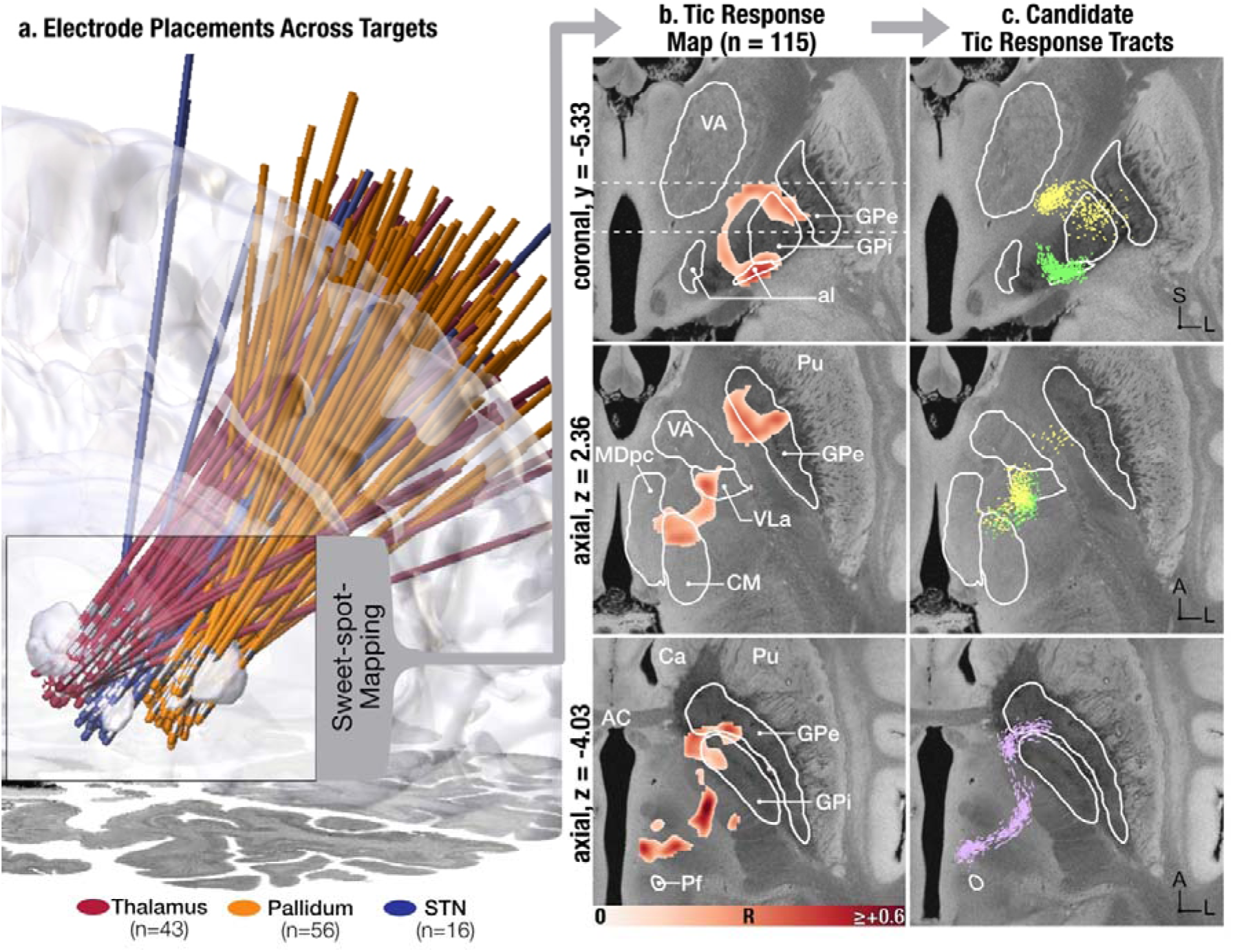
DBS Response Circuitry. **a)** The voxel-wise sweetspot analysis was repeated across the entire cohort (N=115). Electrode localizations are displayed on top of the BigBrain template.^48^ Magenta represents thalamus, orange represents pallidum and blue represents STN cohorts. Centromedian-Parafascicular Complex (CM-Pf, left), subthalamic nucleus (STN, middle) and pallidum are visualized (GPi, GPe, right). **b)** Positive voxels of the tic response map derived from all patients revealed tract-like voxel clusters. This observation can most convincingly be appreciated when scrolling through the maps, as demonstrated in the Supplementary Video. **c)** Based on our *a priori* anatomical tracts and discussions with experts, the sweetspot landscape could be condensed into three anatomical bundles: a subregion of the ansa lenticularis (green) and fasciculus lenticularis (yellow), as well as an outflow bundle projecting from intralaminar nuclei to the lentiform nucleus (purple). A direct comparison to *a priori* tracts (which had been created without knowledge of the sweetspot) is visualized in Supplementary Fig. 14.

While the DBS response (‘sweetspot’) landscape constitutes the final model of optimal placement (as it is built directly from the data and hence represents it best), we tested how much variance in clinical outcomes these three tracts could possibly explain, and whether they would equally explain variance across all three targets. Critically, since the tracts were informed by the sweetspot landscape, the following analyses are circular. The analysis after subjecting tracts to fiber filtering are even more circular. The aim of these results is to quantify the amount of variance that could be possibly explained by a comparably simple model that constituted only of three tracts.

Overlaps between E-fields and the three-tract circuitry (Fig. 4a) was significantly associated with tic outcomes (*R*=0.30, *p*=0.002), which naturally increased further after refining tracts in data-driven fashion by DBS fiber filtering (see methods, *R*=0.55, *p*=3.999e-4, *R^2^* ≈ 0.30). The relationship held true when testing just the thalamus (unweighted tracts (uw): *R*=0.39, *p*=0.013, after fiber-filtering (ff): *R*=0.55, *p*=3.999e-4) and pallidum cohorts (uw: *R*=0.47, *p*=5.999e-4, ff: *R*=0.48, *p*=2.000e-4). The STN cohort correlated with a similar magnitude but the result was not significant given low N (uw: *R*=0.36, *p*=0.223, ff: *R*=0.44, *p*=0.092). Figure 5 shows stimulation sites of individual example patients in synopsis with the tract landscape.

**Figure 4:**
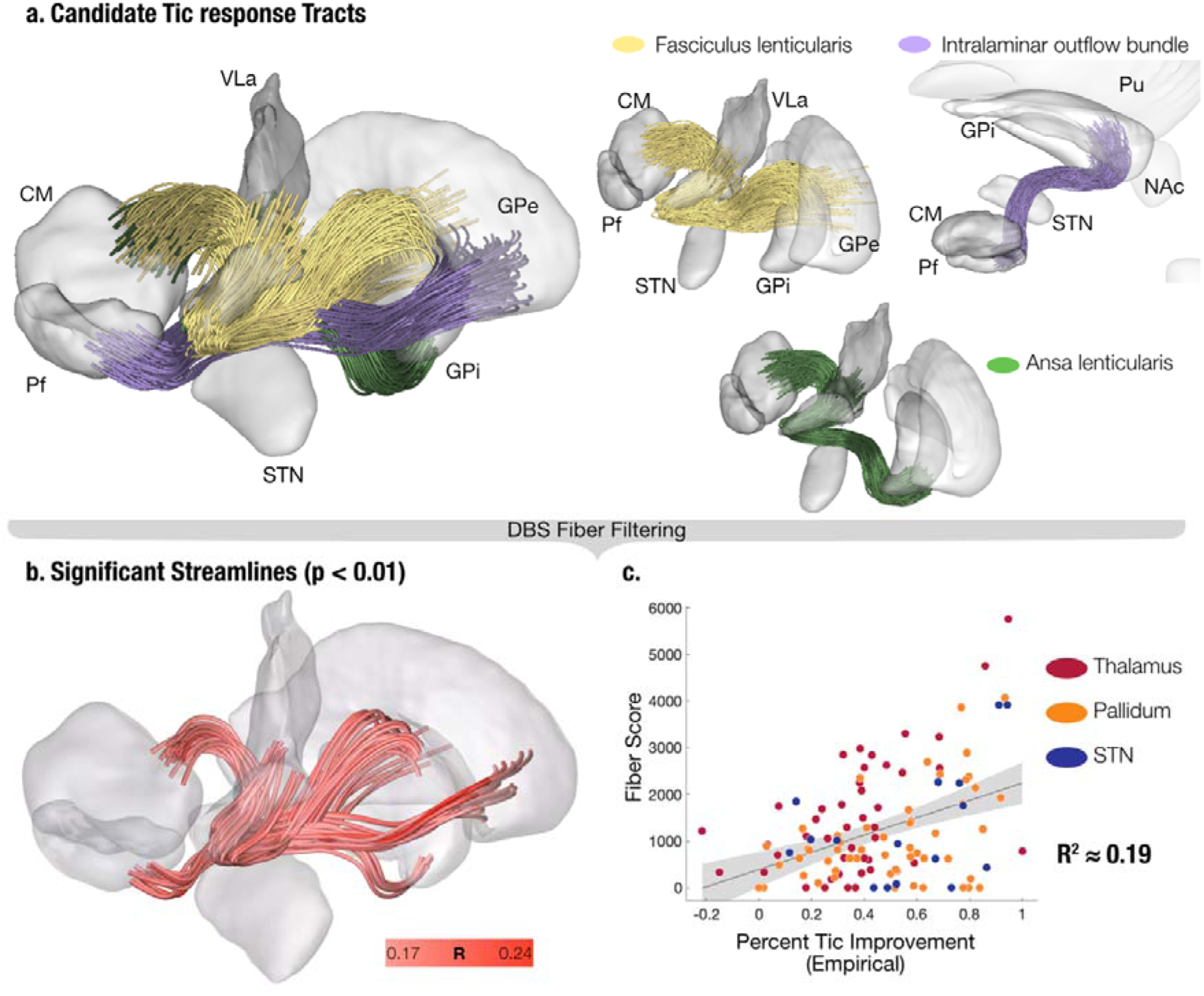
Candidate tic response tracts. **a)** A sagittal 3D view of the candidate tic response tracts. Fasciculus lenticularis (yellow) leaves GPi dorsally, whereas ansa lenticularis (green) exits ventrally, forming two main components of the pallidothalamic pathway. Intralaminar outflow bundle (purple) gives collaterals to limbic parts of GPi/GPe, before reaching their main destination, ventral striatum. **b)** The three tracts were fed into DBS fiber filtering to test stimulation of any streamlines correlate significantly with tic outcome. Streamlines that were not significantly correlated with outcomes were discarded, leaving the general circuitry unchanged. The resulting model explained 19% of the variance in clinical outcomes across the full cohort.

**Figure 5:**
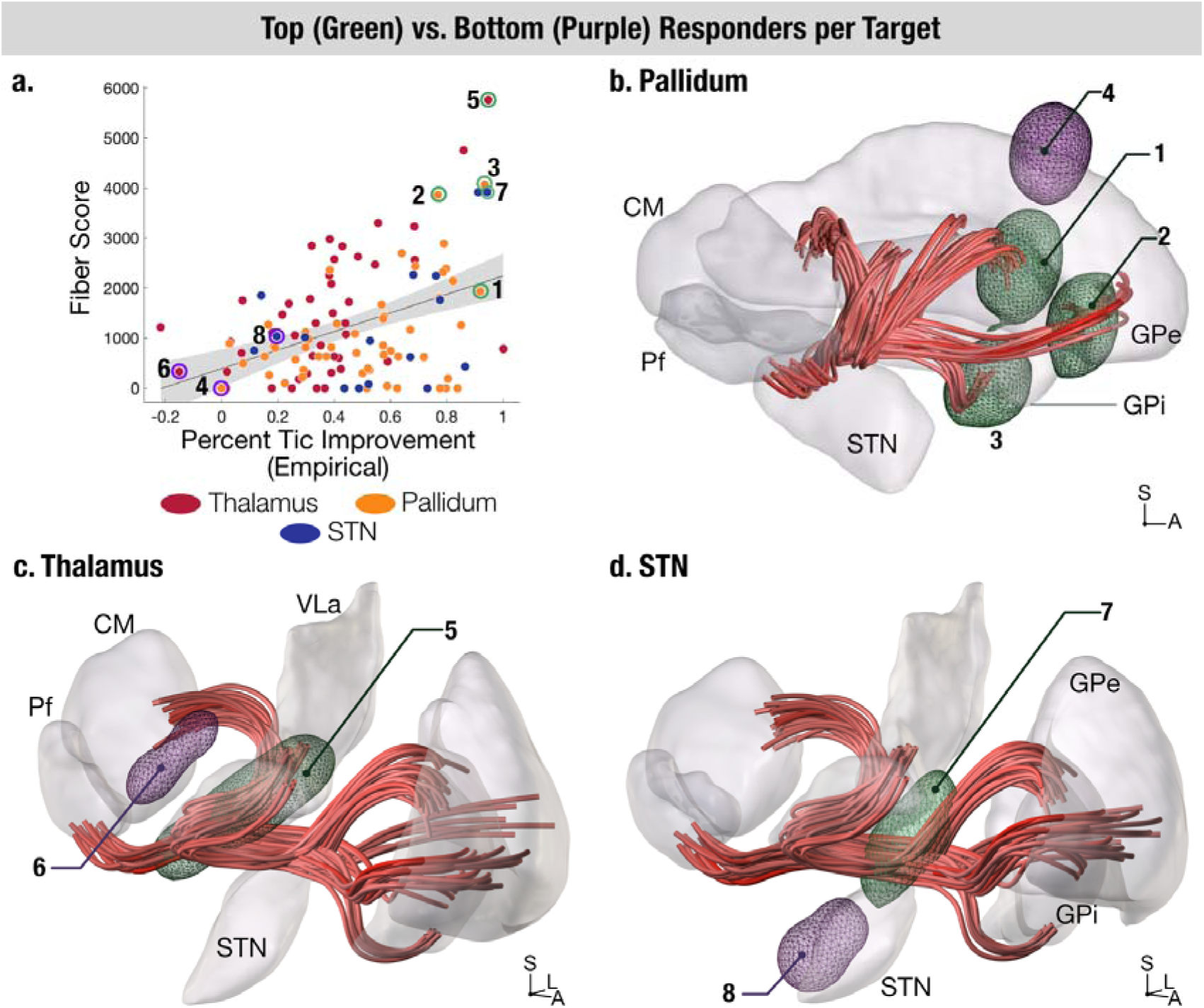
Individual examples. Stimulation fields of top (green) and bottom (purple) responders implanted in each target location are visualized in 3D space for pallidum (b), thalamus (c) and STN (d) are overlaid onto streamlines that survived significance thresholding (p<0.01). Examples from panels b-d) are marked with circles in the scatter plot in panel a), showing their ‘fiber score’ (estimate of the model) and their clinical tic improvement as measured by percentage YGTSS improvements.

**Figure 6:**
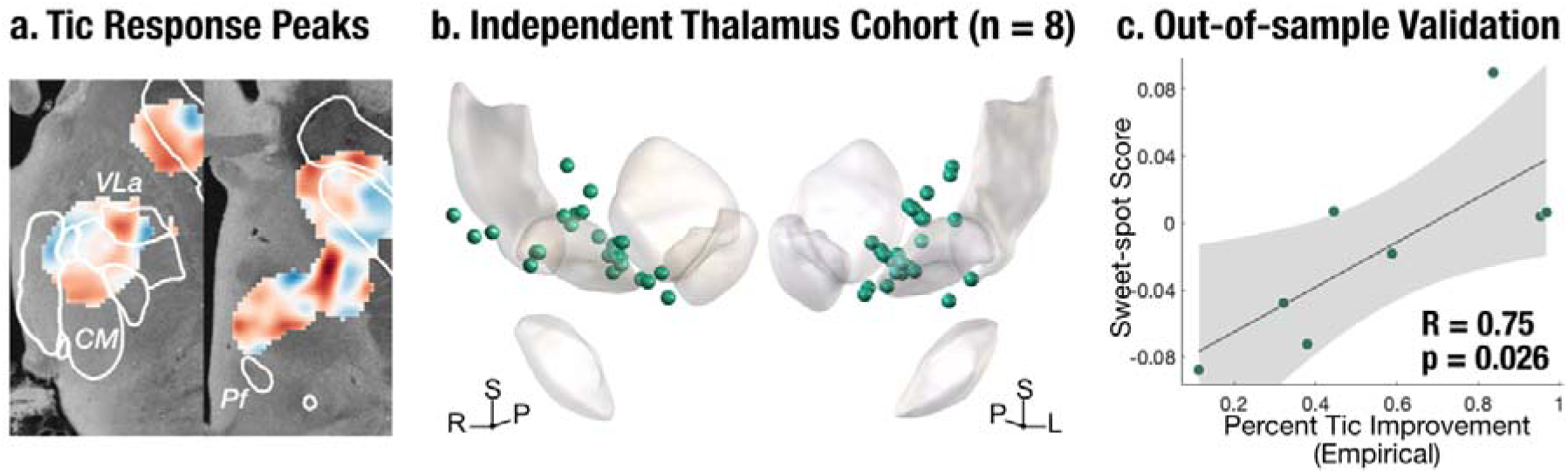
Independent validation of thalamic tic response peaks. Spatial similarity to the tic response map of the full cohort **(a)** was correlated with empirical improvement of an independent cohort implanted in thalamus. **b)** Active contacts of the independent cohort are displayed in 3D in form of spheres. **c)** Empirical tic improvement was significantly associated with the spatial similarity of E-fields to the tic response map.

### Validation Cohort

*N*=8 additional patients operated in Cologne could be post-hoc retrieved from our database that had not been published before, i.e. were entirely independent from the analyses reported thus far. Their mean age at surgery was 25.4 years ± 3.6. Average tic improvement after DBS implantation was 59.3% ± 30.4 at a median follow-up of 16 months [IQR: 10.25-64.25]. (Supplementary Table 1)

Estimates about outcomes in this cohort from the sweetspot model significantly correlated with clinical improvements (*R*=0.75, *p*=0.026). While the cohort is small, given the clear hypothesis of a positive association between sweetspot overlaps and clinical outcomes, even a one-sided test (*p*=0.013) would have been appropriate.

## Discussion

In a large retrospective multi-center GTS-DBS imaging cohort (*N*=115), we investigated the DBS response maps in a pooled analysis across patients targeting thalamus, pallidum and subthalamic nucleus areas. We demonstrated the local tic response hubs within each target, as well as the OCB response hubs within thalamus and pallidum. Tic improvement of the STN patients could be explained by the tic response maps generated from thalamus and pallidum, suggesting a common network. Indeed, tic response maps peaked along known anatomical tracts, revealing a common subcortical network. Anatomically, the shared tic network converged on subcomponents that predominantly mapped to three pathways: the ansa lenticularis, the fasciculus lenticularis and the outflow bundles projecting from intralaminar thalamic nuclei to the lenticular nucleus. This single fiber model explained 19% variance across all initial cohorts (*N*=115) and may generate hypotheses about the structural connections needed to stimulate in DBS for GTS. The final tic response landscape was able to explain significant amounts of variance in an independent patient cohort.

### Local response topography

Voxel-wise correlations revealed three local peaks per target in thalamus and pallidum. (Fig. 1) The mid-ventral (P2)^49^ and anteromedial GPi peaks (P3)^50^ were similar to previously described regions associated with tic improvement. We identified an additional sweet-spot, the mid-dorsal GPi peak (P1). P1 and P2 resided in the mid-GPi, corresponding to the zone between motor and associative pallidum. P3, in contrast, resided in the limbic subdomain of GPi. Laterally adjacent to P3, a strong peak in optimal response was seen within the anteromedial GPe, which could either be part of the intralaminary nuclei outflow bundle collaterals to the GPe or part of the striatopallidofugal fibers traversing through the main axis of the basal ganglia. In the thalamus, T1 and T2 peaks resided in pallidal receiving zones of the thalamus.^51–55^ Previous studies described similar regions in smaller samples.^56,57^ Interestingly, the central parts of the CM-Pf complex correlated negatively with tic improvement, despite their use as a target for GTS-DBS by some centers.^7,58^ Stimulation of rostrodorsal CM (T1) and ventral areas of Pf (T3) were associated with better clinical outcomes. The cohort implanted with the STN as target revealed that the stimulation effect was truly delivered not by the STN proper, but by fibers of passage that traverse the nucleus dorsally, interconnecting pallidum and thalamus. Of note, this does not automatically render the STN an invalid target choice. Given its clear visibility on T2-weighted images makes the structure straightforward and safe to target.^9^ Moreover, since the tracts pass a structural bottleneck in its direct dorsal adjacency, it seems like a sensible strategy to target the STN in conventional fashion but place electrodes more dorsally than in Parkinson’s Disease. Similar conclusions have been reached for cervical types of dystonia^59^. However, potentially, one should speak of DBS to the Fields of Forel^60^ instead of STN-DBS for GTS, in the future.

For OCB, the pallidal peak (*n*=22) partly overlapped with the dorsal tic peak (P1) and extended rostrally (Supplementary Fig. 9). This was consistent with previous reports^12^ and is expected considering the significant correlation between tic and OCB outcomes in pallidum. Furthermore, a recent trans-diagnostic study showed that in STN-DBS, clinical response for obsessive-compulsive disorder was associated with stimulating more anteriorly when compared to GTS^20^. Our results may reflect the same or a similar gradient in the pallidum. Our findings were in line (and partly overlapping) with previous reports of better OCB improvement in pallidal when compared to thalamic DBS for GTS^7^, with only three thalamic patients improving on the Y-BOCS. One reason could be that the thalamic OCB peak (*n*=16) spatially diverged from the thalamic tic response peak. This result should be interpreted with caution, since the average OCB improvement in thalamic cohort was ∼zero and the peak might have been dominated by the three mentioned patients who showed >15% improvement. Critically, however, positive correlations with Y-BOCS improvements could also be driven by stimulation sites associated with less worsening, so the peak is not exclusively informed by just three data points. We would still argue that, based on this and prior evidence^7^, the pallidum would serve as a better target in the light of high OCB since i) it is more effective to treat OCB and ii) targets of optimal tic and OCB response overlap spatially and can hence be reached by the same electrode.

Previous attempts to map local response peaks in GTS-DBS were limited by small sample sizes^49,50,56,57^. A previous study of 63 individuals was inconclusive largely as a result of a large overlap of responder and non-responder VTAs within the target structures.^12^ A structural connectivity based large-scale study later identified lateral CM and the region anterolateral to the nucleus as a peak region by inverse mapping of optimal structural connectivity^23^. This region broadly corresponds to peaks seen here, especially with the outflow bundle that sends projections from intralaminar nuclei to the lentiform nucleus. Our study applied a recently introduced E-field based sweetspot mapping method, which may be more sensitive to fine-grained anatomy since stimulation volumes are not modeled in binary (all-or-nothing) fashion^38^. Moreover, the method facilitates delineation of symptom response landscapes accounting for uncertainty in neural activation thresholds and response curves^38,39,61^.

Of note, the present study focused on the optimal stimulation sites within each target region and is not designed to compare clinical efficiency across targets. Only the Y-BOCS scores were compared given the stark difference and to assess the interpretability of the thalamic OCB response map (Supplementary Figs. 10-12).

### From Voxels to Networks

While local maps highlighted optimal target sites, specific pathways possibly mediating this treatment response would be helpful to enhance our mechanistic understanding.^7,9^ Our next analyses uncovered a common circuitry of specific subcomponents of ansa lenticularis, fasciculus lenticularis and thalamic projections connecting the intralaminar thalamic nuclei to the lentiform nucleus^55,62,63^ (Figs. 3,4, Supplementary Video). Two of these anatomical components, the ansa lenticularis and the lenticular fasciculus have been previously characterized as a functional continuum.^17,52^ The pallidothalamic component of the proposed tic-response circuitry emerged as a single entity located at the border of associative and motor GPi, entering the supplementary motor area projecting zones (VLa)^64^ of thalamus. The thalamic outflow bundle, likely originating from Pf or medial CM, projected back to the striatum, GPe and GPi, predominantly using the limbic regions.

Evidence from prior work has already implicated all of these regions in the production and suppression of tics. First, electrophysiology studies support the shared network hypothesis, demonstrated by low-frequency coupling between CM and GPi as recorded from DBS electrodes in humans^14,15^. Additionally, a previous cross-target study (*n*=21) identified the broader ventral pallidofugal pathway, inclusive of the ansa lenticularis^49^. The International GTS-DBS Registry identified a similar sub-bundle as the most commonly activated within fasciculus lenticularis^65^. Moreover, increased connectivity between thalamus and putamen in GTS patients^66^, convergence of networks involved in tic-inducing lesions and GTS-DBS to anterior putamen^21^ and increased striatal dopamine levels after CM-Pf stimulation associating with tic improvement in rats^67^ all collectively support the involvement of the wider efferent zone of a thalamic outflow bundle.

### Limitations

Our study has several important limitations. The proposed network model was built in exploratory and circular fashion since it was based on sweetspots derived from the same retrospective cohort it was tested on. This renders our three-tract model as a hypothesis-generating, rather than -confirming result, i.e. the tracts, which we make openly available, should be validated by additional datasets or, optimally, within prospective trials. Despite this limitation, the model, composed of three simple anatomical bundles of which two are a functional continuum, was able to account for clinical outcome variance across a large cohort and across three target sites. Detailed symptom topography was not available. It is likely that distinct tic subtypes may engage different subcircuits, similar to symptom-specific patterns described in Parkinson’s disease.^68^ We opine that it may be possible in the future to further personalize brain targeting for GTS-DBS, if similar studies are repeated for clusters of phenotypes that are more similar to one another (e.g. vocal vs. motor tics). Specific components of the described circuitry may be more relevant to different forms of tics, but we were unable to test this based on the data at hand. Despite the use of a state-of-the-art dedicated DBS modeling pipeline, methodological factors such as assumptions of E-field modeling as well as spatial inaccuracies due to image coregistrations could have introduced bias and affected the precision of the results. Finally, our independent test cohort was composed of patients implanted in thalamus only and came from the same center (Cologne) as some of the initial patients (*n*=13 out of 43 patients). Future investigations on thalamic cohorts from additional centers and targeting the pallidal and subthalamic region are necessary to further assess the generalizability of our results.

## Conclusions

We conclude that optimal DBS target structures are embodied by a three-fold tract landscape that interconnects pallidum and thalamus. Namely, three specific bundles – subcomponents of ansa lenticularis, fasciculus lenticularis and efferents from intralaminary thalamic nuclei to the lentiform nucleus – were functionally and anatomically important to the GTS-DBS clinical response. The findings should be useful for neurosurgeons targeting and healthcare providers programming GTS DBS.

## Data Availability

All data produced in the present study are available upon reasonable request to the authors.

## Acknowledgements

In-depth discussions about the anatomical correlates of presented sweetspot models were held with multiple international expert anatomists of the subthalamic region including co-authors A. S., M. P. as well as Jürgen K. Mai, whom the authors would like to thank.

## Author Contributions

I.A.S.: conceptualization, data curation, methodology, formal analysis, visualization, writing – original draft, and funding acquisition. K.B.: methodology, software, formal analysis, supervision (supporting), resources, writing – review and editing. K.A.J: data curation, formal analysis, writing – review and editing. H.F.: investigation, resources, writing – review and editing. S.O.: methodology, software, writing – review and editing. N.L.: methodology, software, supervision (supporting), resources, writing – review and editing. B.H., L.G.G.: data curation, formal analysis, writing – review and editing. P.Z., G.M.M., N.R., C.N., M.M.: data curation, methodology, software, formal analysis, writing – review and editing. A.T., W.J.N.: resources, supervision (supporting), writing – review and editing. T.D.: conceptualization, investigation, formal analysis, resources, writing – review and editing. A.C.v.O.: visualization, writing – review and editing. R.S.: data curation, writing – review and editing. J.C.B., C.v.d.L, J.K., D.H., P.A., R.S., P.H., Y.W., N.P., C.K., M.L.W., L.A., A.Y.J.M., A.F.G.L., C.Z., B.S. J.G.Z., F.G.M., X.Y., X.X., Z.L., D.S., A.B., M.P., A.Y.M., M.H.P., J.L.O., T.F., P.L., L.Z., E.M.J., Z.K., A.A.P., L.M.R., M.V., A.M., O.V.F., A.M.L., K.D.F., C.R.B., V.V.V., M.S.O.: investigation, resources, data curation, writing – review and editing. M.P., A.F.S.: formal analysis, writing – review and editing. A.H.: conceptualization, data curation, methodology, software, resources, formal analysis, visualization, writing – original draft, review and editing, supervision (lead), project administration and funding acquisition.

## References

1. Johnson KA, Worbe Y, Foote KD, Butson CR, Gunduz A, Okun MS. Tourette syndrome: clinical features, pathophysiology, and treatment. The Lancet Neurology. 2023;22(2):147–158. doi:10.1016/S1474-4422(22)00303-9

2. Szejko N, Worbe Y, Hartmann A, et al. European clinical guidelines for Tourette syndrome and other tic disorders—version 2.0. Part IV: deep brain stimulation. Eur Child Adolesc Psychiatry. 2022;31(3):443–461. doi:10.1007/s00787-021-01881-9

3. Billnitzer A, Jankovic J. Current Management of Tics and Tourette Syndrome: Behavioral, Pharmacologic, and Surgical Treatments. Neurotherapeutics. 2020;17(4):1681–1693. doi:10.1007/s13311-020-00914-6

4. Vandewalle V, Linden C van der, Groenewegen HJ, Caemaert J. Stereotactic treatment of Gilles de la Tourette syndrome by high frequency stimulation of thalamus. The Lancet. 1999;353(9154):724. doi:10.1016/S0140-6736(98)05964-9

5. Rickards H, Wood C, Cavanna AE. Hassler and Dieckmann’s seminal paper on stereotactic thalamotomy for Gilles de la Tourette syndrome: Translation and critical reappraisal. Movement Disorders. 2008;23(14):1966–1972. doi:10.1002/mds.22238

6. Visser-Vandewalle V, Temel Y, Boon P, et al. Chronic bilateral thalamic stimulation: a new therapeutic approach in intractable Tourette syndrome. Published online December 1, 2003. doi:10.3171/jns.2003.99.6.1094

7. Wehmeyer L, Schüller T, Kiess J, et al. Target-Specific Effects of Deep Brain Stimulation for Tourette Syndrome: A Systematic Review and Meta-Analysis. Frontiers in Neurology. 2021;12. Accessed November 4, 2022. https://www.frontiersin.org/articles/10.3389/fneur.2021.769275

8. Vilela-Filho O, Souza JT, Ragazzo PC, et al. Bilateral Globus Pallidus Externus Deep Brain Stimulation for the Treatment of Refractory Tourette Syndrome: An Open Clinical Trial. Neuromodulation. 2023;0(0). doi:10.1016/j.neurom.2023.04.473

9. Dai L, Xu W, Song Y, et al. Subthalamic deep brain stimulation for refractory Gilles de la Tourette’s syndrome: clinical outcome and functional connectivity. J Neurol. 2022;269(11):6116–6126. doi:10.1007/s00415-022-11266-w

10. Vissani M, Cordella R, Micera S, Eleopra R, Romito LM, Mazzoni A. Spatio-temporal structure of single neuron subthalamic activity identifies DBS target for anesthetized Tourette syndrome patients. J Neural Eng. 2019;16(6):066011. doi:10.1088/1741-2552/ab37b4

11. Neudorfer C, El Majdoub F, Hunsche S, Richter K, Sturm V, Maarouf M. Deep Brain Stimulation of the H Fields of Forel Alleviates Tics in Tourette Syndrome. Front Hum Neurosci. 2017;11. doi:10.3389/fnhum.2017.00308

12. Johnson KA, Fletcher PT, Servello D, et al. Image-based analysis and long-term clinical outcomes of deep brain stimulation for Tourette syndrome: a multisite study. J Neurol Neurosurg Psychiatry. 2019;90(10):1078–1090. doi:10.1136/jnnp-2019-320379

13. Singer HS, Pellicciotti J. The Pathophysiology of Tics: An Anatomic Review. Psychiatric Clinics of North America. 2025;48(1):15–29. doi:10.1016/j.psc.2024.08.003

14. Lowor G, Gomez J, Hook M, et al. The CM-aGPi Network in the Generation of Tics in Tourette Syndrome. Mov Disord. Published online September 22, 2025. doi:10.1002/mds.70043

15. Neumann WJ, Huebl J, Brücke C, et al. Pallidal and thalamic neural oscillatory patterns in tourette’s syndrome. Annals of Neurology. 2018;84(4):505–514. doi:10.1002/ana.25311

16. Nieuwenhuys R, Voogd J, Huijzen C van. The Human Central Nervous System. Fourth Edition. Springer-Verlag Berlin Heidelberg New York; 2008.

17. Neudorfer C, Maarouf M. Neuroanatomical background and functional considerations for stereotactic interventions in the H fields of Forel. Brain Struct Funct. 2018;223(1):17–30. doi:10.1007/s00429-017-1570-4

18. Li N, Baldermann JC, Kibleur A, et al. A unified connectomic target for deep brain stimulation in obsessive-compulsive disorder. Nature Communications. 2020;11(1). doi:10.1038/s41467-020-16734-3

19. Sobesky L, Goede L, Odekerken VJJ, et al. Subthalamic and pallidal deep brain stimulation: are we modulating the same network? Brain. 2022;145(1):251–262. doi:10.1093/brain/awab258

20. Hollunder B, Ostrem JL, Sahin IA, et al. Mapping dysfunctional circuits in the frontal cortex using deep brain stimulation. Nat Neurosci. 2024;27(3):573–586. doi:10.1038/s41593-024-01570-1

21. Ganos C, Al-Fatly B, Fischer JF, et al. A neural network for tics: insights from causal brain lesions and deep brain stimulation. Brain. Published online January 13, 2022:awac009. doi:10.1093/brain/awac009

22. Baldermann JC, Hennen C, Schüller T, et al. Normative Functional Connectivity of Thalamic Stimulation for Reducing Tic Severity in Tourette Syndrome. Biological Psychiatry: Cognitive Neuroscience and Neuroimaging. 2022;7(8):841–844. doi:10.1016/j.bpsc.2022.01.009

23. Johnson KA, Duffley G, Anderson DN, et al. Structural connectivity predicts clinical outcomes of deep brain stimulation for Tourette syndrome. Brain. 2020;143(8):2607–2623. doi:10.1093/brain/awaa188

24. Leckman JF, Riddle MA, Hardin MT, et al. The Yale Global Tic Severity Scale: Initial Testing of a Clinician-Rated Scale of Tic Severity. Journal of the American Academy of Child & Adolescent Psychiatry. 1989;28(4):566–573. doi:10.1097/00004583-198907000-00015

25. Goodman WK, Price LH, Rasmussen SA, et al. The Yale-Brown Obsessive Compulsive Scale: I. Development, Use, and Reliability. Arch Gen Psychiatry. 1989;46(11):1006–1011. doi:10.1001/archpsyc.1989.01810110048007

26. Neudorfer C, Butenko K, Oxenford S, et al. Lead-DBS v3.0: Mapping deep brain stimulation effects to local anatomy and global networks. NeuroImage. 2023;268:119862. doi:10.1016/j.neuroimage.2023.119862

27. Avants BB, Tustison NJ, Song G, Cook PA, Klein A, Gee JC. A reproducible evaluation of ANTs similarity metric performance in brain image registration. NeuroImage. 2011;54(3):2033–2044. doi:10.1016/j.neuroimage.2010.09.025

28. Avants BB, Epstein CL, Grossman M, Gee JC. Symmetric diffeomorphic image registration with cross-correlation: Evaluating automated labeling of elderly and neurodegenerative brain. Medical Image Analysis. 2008;12(1):26–41. doi:10.1016/j.media.2007.06.004

29. Friston KJ, Holmes AP, Worsley KJ, Poline JP, Frith CD, Frackowiak RSJ. Statistical parametric maps in functional imaging: A general linear approach. Human Brain Mapping. 1994;2(4):189–210. doi:10.1002/hbm.460020402

30. Fonov V, Evans A, McKinstry R, Almli C, Collins D. Unbiased nonlinear average age-appropriate brain templates from birth to adulthood. NeuroImage. 2009;47:S102. doi:10.1016/S1053-8119(09)70884-5

31. Iglesias JE. A ready-to-use machine learning tool for symmetric multi-modality registration of brain MRI. Sci Rep. 2023;13(1):6657. doi:10.1038/s41598-023-33781-0

32. Hoffmann M, Billot B, Greve DN, Iglesias JE, Fischl B, Dalca AV. SynthMorph: Learning Contrast-Invariant Registration Without Acquired Images. IEEE Transactions on Medical Imaging. 2022;41(3):543–558. doi:10.1109/TMI.2021.3116879

33. Oxenford S, Ríos AS, Hollunder B, et al. WarpDrive: Improving spatial normalization using manual refinements. Medical Image Analysis. 2024;91:103041. doi:10.1016/j.media.2023.103041

34. Husch A, V. Petersen M, Gemmar P, Goncalves J, Hertel F. PaCER - A fully automated method for electrode trajectory and contact reconstruction in deep brain stimulation. NeuroImage: Clinical. 2018;17:80–89. doi:10.1016/j.nicl.2017.10.004

35. Horn A, Kühn AA. Lead-DBS: A toolbox for deep brain stimulation electrode localizations and visualizations. NeuroImage. 2015;107:127–135. doi:10.1016/j.neuroimage.2014.12.002

36. Billot B, Greve DN, Puonti O, et al. SynthSeg: Segmentation of brain MRI scans of any contrast and resolution without retraining. Med Image Anal. 2023;86:102789. doi:10.1016/j.media.2023.102789

37. Butenko K, Bahls C, Schröder M, Köhling R, van Rienen U. OSS-DBS: Open-source simulation platform for deep brain stimulation with a comprehensive automated modeling. PLoS Comput Biol. 2020;16(7):e1008023. doi:10.1371/journal.pcbi.1008023

38. Horn A, Reich MM, Ewert S, et al. Optimal deep brain stimulation sites and networks for cervical vs. generalized dystonia. Proc Natl Acad Sci USA. 2022;119(14):e2114985119. doi:10.1073/pnas.2114985119

39. Ríos AS, Oxenford S, Neudorfer C, et al. Optimal deep brain stimulation sites and networks for stimulation of the fornix in Alzheimer’s disease. Nat Commun. 2022;13(1):7707. doi:10.1038/s41467-022-34510-3

40. Åström M, Diczfalusy E, Martens H, Wårdell K. Relationship between neural activation and electric field distribution during deep brain stimulation. IEEE Trans Biomed Eng. 2015;62(2):664–672. doi:10.1109/TBME.2014.2363494

41. Irmen F, Horn A, Mosley P, et al. Left Prefrontal Connectivity Links Subthalamic Stimulation with Depressive Symptoms. Annals of Neurology. 2020;87(6):962–975. doi:10.1002/ana.25734

42. Hart LA, Warren AEL, Pacheco-Barrios N, et al. Deep Brain Stimulation for Epilepsy: Optimal Targeting and Clinical Outcomes. medRxiv. Preprint posted online June 23, 2025:2025.06.23.25330152. doi:10.1101/2025.06.23.25330152

43. Cacciatore M, Magnani FG, Barbadoro F, et al. Thalamus and consciousness: a systematic review on thalamic nuclei associated with consciousness. Front Neurol. 2025;16:1509668. doi:10.3389/fneur.2025.1509668

44. Friedrich H, Sahin AI, Rajamani N, et al. A precise atlas of the human subcortex. bioRxiv. Preprint posted online February 15, 2026:2026.02.13.705755. doi:10.64898/2026.02.13.705755

45. Kikinis R, Pieper SD, Vosburgh KG. 3D Slicer: A Platform for Subject-Specific Image Analysis, Visualization, and Clinical Support. In: Jolesz FA, ed. Intraoperative Imaging and Image-Guided Therapy. Springer; 2014:277–289. doi:10.1007/978-1-4614-7657-3_19

46. Edlow BL, Mareyam A, Horn A, et al. 7 Tesla MRI of the ex vivo human brain at 100 micron resolution. Sci Data. 2019;6(1):244. doi:10.1038/s41597-019-0254-8

47. Chua MMJ, Pinzon AM, Neudorfer C, et al. Optimal focused ultrasound lesion location in essential tremor. Sci Adv. 2025;11(20):eadp0532. doi:10.1126/sciadv.adp0532

48. Amunts K, Lepage C, Borgeat L, et al. BigBrain: An Ultrahigh-Resolution 3D Human Brain Model. Science. 2013;340(6139):1472–1475. doi:10.1126/science.1235381

49. Avecillas-Chasin JM, Galbiati T, Porta M, Servello D. Deep brain stimulation for Tourette syndrome: modulation of the limbic-motor interface network. Journal of Neurosurgery. 2023;1(aop):1–10. doi:10.3171/2023.10.JNS231317

50. Akbarian-Tefaghi L, Akram H, Johansson J, et al. Refining the Deep Brain Stimulation Target within the Limbic Globus Pallidus Internus for Tourette Syndrome. Stereotactic and Functional Neurosurgery. 2017;95(4):251–258. doi:10.1159/000478273

51. Neudorfer C, Kultas-Ilinsky K, Ilinsky I, et al. The role of the motor thalamus in deep brain stimulation for essential tremor. Neurotherapeutics. 2024;21(3):e00313. doi:10.1016/j.neurot.2023.e00313

52. Parent M, Parent A. The pallidofugal motor fiber system in primates. Parkinsonism & Related Disorders. 2004;10(4):203–211. doi:10.1016/j.parkreldis.2004.02.007

53. Sakai ST, Inase M, Tanji J. Comparison of cerebellothalamic and pallidothalamic projections in the monkey (Macaca fuscata): A double anterograde labeling study. Journal of Comparative Neurology. 1996;368(2):215–228. doi:10.1002/(SICI)1096-9861(19960429)368:2%3C215::AID-CNE4%3E3.0.CO;2-6

54. Mehler WR, Nauta WJ. Connections of the basal ganglia and of the cerebellum. Confin Neurol. 1974;36(4-6):205–222. doi:10.1159/000102797

55. Sadikot AF, Rymar VV. The primate centromedian–parafascicular complex: Anatomical organization with a note on neuromodulation. Brain Research Bulletin. 2009;78(2):122–130. 10.1016/j.brainresbull.2008.09.016

56. Baldermann JC, Kuhn J, Schüller T, et al. Thalamic deep brain stimulation for Tourette Syndrome: A naturalistic trial with brief randomized, double-blinded sham-controlled periods. Brain Stimulation. 2021;14(5):1059–1067. doi:10.1016/j.brs.2021.07.003

57. Morishita T, Sakai Y, Iida H, et al. Neuroanatomical considerations for optimizing thalamic deep brain stimulation in Tourette syndrome. Journal of Neurosurgery. 2021;136(1):231–241. doi:10.3171/2021.2.JNS204026

58. Aydin S, Darko K, Jenkins A, Detchou D, Barrie U. Deep brain stimulation for Tourette’s syndrome. Neurosurg Rev. 2024;47(1):734. doi:10.1007/s10143-024-02958-0

59. Butenko K, Neudorfer C, Dembek TA, et al. Engaging dystonia networks with subthalamic stimulation. Proc Natl Acad Sci USA. 2025;122(2):e2417617122. doi:10.1073/pnas.2417617122

60. Horisawa S, Kohara K, Murakami M, Fukui A, Kawamata T, Taira T. Deep Brain Stimulation of the Forel’s Field for Dystonia: Preliminary Results. Front Hum Neurosci. 2021;15:768057. doi:10.3389/fnhum.2021.768057

61. Butenko K, Roediger J, Al-Fatly B, et al. Activation metrics for structural connectivity recruitment in deep brain stimulation. Brain Communications. 2025;7(5):fcaf301. doi:10.1093/braincomms/fcaf301

62. Parent M, Parent A. Single-axon tracing and three-dimensional reconstruction of centre médian-parafascicular thalamic neurons in primates. Journal of Comparative Neurology. 2005;481(1):127–144. doi:10.1002/cne.20348

63. Sadikot AF, Parent A, François C. Efferent connections of the centromedian and parafascicular thalamic nuclei in the squirrel monkey: A PHA-L study of subcortical projections. Journal of Comparative Neurology. 1992;315(2):137–159. doi:10.1002/cne.903150203

64. Haber SN, Calzavara R. The cortico-basal ganglia integrative network: the role of the thalamus. Brain Res Bull. 2009;78(2-3):69–74. doi:10.1016/j.brainresbull.2008.09.013

65. Johnson KA, Duffley G, Foltynie T, et al. Basal Ganglia Pathways Associated With Therapeutic Pallidal Deep Brain Stimulation for Tourette Syndrome. Biological Psychiatry: Cognitive Neuroscience and Neuroimaging. 2021;6(10):961–972. 10.1016/j.bpsc.2020.11.005

66. Worbe Y, Marrakchi-Kacem L, Lecomte S, et al. Altered structural connectivity of cortico-striato-pallido-thalamic networks in Gilles de la Tourette syndrome. Brain. 2015;138(Pt 2):472–482. doi:10.1093/brain/awu311

67. Rusheen AE, Rojas-Cabrera J, Goyal A, et al. Deep brain stimulation alleviates tics in Tourette syndrome via striatal dopamine transmission. Brain. 2023;146(10):4174–4190. doi:10.1093/brain/awad142

68. Rajamani N, Friedrich H, Butenko K, et al. Deep brain stimulation of symptom-specific networks in Parkinson’s disease. Nat Commun. 2024;15(1):4662. doi:10.1038/s41467-024-48731-1

