## Supplementary Material for "Optimal Deep Brain Stimulation Locations for Gilles de la Tourette Syndrome"

Sahin *et al.*

**Contents**

**Page**

**Supplementary Methods	2**

Dataset Description	2

**Fig S1:**	Flowchart of the inclusion-exclusion process	2

**Table S1:**	Patient demographics and clinical Information	3-6

Anatomical Pathway Modelling	7

**Fig S2:**	Delineation of a-priori tract candidates led to a 3D subcortical DBS	8

atlas in stereotactic standard space.

**Table S2:**	List of pathways in the *a priori* subcortical white matter atlas	10-11

Example tract modeling steps	12

**Fig S3:**	Striatopallidofugal pathways	13

**Fig S4:**	Efferents of centromedian Nucleus	14

**Fig S5:**	Efferents of parafascicular Nucleus	15

**Fig S6:**	Organization of thalamic afferents	16

**Fig S7:**	Cerebellar vs. pallidal afferent zones of thalamus	16

**Fig S8:**	Pallidothalamic pathways	17

**Fig S9:**	Cerebellothalamic pathways	18

**Supplementary Results	19**

**Fig S10:**	Tic vs. obsessive compulsive behavior improvement across cohorts	19

**Fig S11:**	Tic vs. obsessive compulsive behavior response maps	20

**Fig S12:**	Top vs. Bottom OCB Responders from the thalamus cohort	21

**Fig S13:**	Tic response peaks for the subthalamic nucleus	22

**Fig S14:**	Sweetspot data, a-priori anatomical tracts and candidate response tracts	23

**Supplementary References	24**

##### **Tourette-DBS: Dataset Description**

The dataset for this study consisted of a total of 115 patients who had undergone bilateral DBS treatment for Tourette Syndrome in 12 different centers world-wide. Imaging and clinical data was compiled for 15 patients from Cologne^1^, 6 patients from Maastricht^1^, 9 patients from Paris^1^, 4 patients from Milan/Pisa, 10 patients from Shanghai^2^, 7 patients from Brazil^3^ and 64 patients from International Tourette Syndrome DBS Database and Registry^4^. Inclusion/exclusion process is illustrated as a flowchart in supplementary Figure 1.

##### **Supplementary Figure 1: Flowchart of the Inclusion-Exclusion Process**

Retrospective unpublished data from host institution

N=8

Initial Cohort

Patients included in the analysis
N=115

Data included in Johnson et al. (N=64)
Data included in Ganos et al. 2022 (N=30)
Data included in Hollunder et al. (N=14)
Data included in Vilelo-Filho et al. (N=7)

Tic Response Landscape
N=115

OCB Response Landscape
N=38

Incomplete/Unavailable Assessments (n=77)

Validation Cohort

Patients included for validation of the thalamic tic response map

N=8

#### **Supplementary Table 1: Patient Demographics and Clinical Information**

| **Target** | **Thalamus** | | | | | | | |
| --- | --- | --- | --- | --- | --- | --- | --- | --- |
| **Cohort** | Cologne | Maastricht | Italy | NYU | UFL | UCSF | **Thalamus full cohort** | |
| Surgical center | University Hospital Cologne | Maastricht University Medical Center | IRCCS Istituto Ortopedico Galeazzi, Milan, Italy | New York University Medical Center | University of Florida, Gainesville | University of California San Francisco |  | |
| Target | CM/Voi | CM/Pf | CM/Pf | CM/Pf | CM/Pf | CM/Pf |  | |
| Number of patients | 13 | 3 | 7 | 12 | 7 | 1* | 43 | |
| Median Time at FU (months, IQR) | 6 | 6 | 6 | 9.5 | 6 | * | 6 [6-6.7] | |
| Related citation | ^1,5^ | ^6^ | ^6^ | ^6^ | ^6^ | ^6^ |  | |
| **Imaging and electrode specifications** | | |  |  |  |  |  | |
| Postoperative imaging modality | CT (n=13) | CT (n=3) | MRI (n=7) | CT (n=12) | CT (n=7) | MRI (n=1) | CT (n=35), MRI (n=8) | |
| Electrode models | MDT 3387 (n=3), MDT 3389 (n=10) | MDT 3387 (n=3) | MDT 3389 (n=7) | MDT 3387 (n=12) | MDT 3387 (n=2), NP DLL-344-3.5 (n=5) | MDT 3387 (n=1) | MDT 3387 (n=21), MDT 3389 (n=17), NP DL-344-3.5 (n=5) | |
| **Demographics** | |  |  |  |  |  |  |  |
| Sex (F/M) | 3F/10M | 0F/3M | 2F/5M | 3F/9M | 5F/2M | * | 13F/30M | |
| Age at surgery (mean ± SD, in years) | 30.5 ± 10.6 | 42.7 ± 6.8 | 34.6 ± 6.4 | 22.1 ± 5.2 | 32.6 ± 4.9 | * | 29.6 ± 9.5 | |
| **Clinical symptom assessments** | |  |  |  |  |  |  |  |
| **Tic Score** | TTSS (/50) | TTSS (/50) | YGTSS (/100) | YGTSS (/100) | YGTSS (/100) | YGTSS (/100) | TTSS (/50) | YGTSS (/100) |
| Baseline (mean ± SD) | 39.5 ± 8.7 | 42.3 ± 4.0 | 55.7 ± 22.7 | 82.8 ± 15.1 | 89.6 ± 8.8 | * | 40.1 ± 8.0 | 77.6 ± 20.4 |
| DBS ON (mean ± SD) | 22.3 ± 7.2 | 20.7 ± 16.7 | 35.6 ± 14.9 | 49.6 ± 26.0 | 68.0 ± 15.7 | * | 22.0 ± 8.9 | 52.6 ± 25.0 |
| Improvement (%) (mean ± SD) | 43.9 ± 10.3 | 53.5 ± 36.4 | 36.1 ± 10.3 | 36.5 ± 36.2 | 24.5 ± 12.4 | -15.1 ± 0 | 36.7 ± 24.5 | |
| **YBOCS** |  |  |  |  |  |  |  |  |
| Available Observations | N/A | N/A | n = 6 | n = 4 | n = 5 | n = 1 | 16 | |
| Baseline (mean ± SD) |  |  | 20.3 ± 6.2 | 11.5 ± 7.1 | 19.2 ± 7.4 | * | 17.9 ± 7.3 | |
| FU (mean ± SD) |  |  | 19.3 ± 6.3 | 9.5 ± 6.7 | 20.8 ± 7.8 | * | 17.8 ± 8.1 | |
| Improvement (%) |  |  | 5.3 ± 9.6 | 5.7 ± 86.6 | -8.5 ± 11.9 | -13.0 | -0.1 ± 40.3 | |

| **Target** | **Pallidum** | | | | | | | | | |
| --- | --- | --- | --- | --- | --- | --- | --- | --- | --- | --- |
| **Cohort** | Maastricht | Paris | Milan/Pisa | Brazil | China | Italy | UCL | UFL | **Pallidum full cohort** | |
| Surgical center | Maastricht University Medical Center | Sorbonne University | Fondazione IRCCS Instituto Neurologico Carlo Besta Milan | Federal University of Goiás, Brazil | Beijing Neurosurgical Institute, China | IRCCS Istituto Ortopedico Galeazzi, Milan, Italy | University College London, United Kingdom | University of Florida, Gainesville |  |  |
| Target | amGPi | amGPi | GPi/GPe | GPe | pvGPi | amGPi | amGPi | pvGPi |  |  |
| Number of patients | 7 | 11 | 2 | 7 | 4 | 9 | 15 | 1* | 56 | |
| Median Time at FU (months) | 13 | N/A | 6 | 6 | 6 | 6 | 9 | * | 6 [6-12] | |
| Related citation | ^1^ | ^1^ | ^6^ | ^3^ | ^6^ | ^6^ | ^6^ | ^6^ |  |  |
| **Imaging and electrode specifications** | | |  |  |  |  |  |  |  |  |
| Postoperative imaging modality | CT (n=3), MRI (n=4) | CT (n=11) | CT (n=2) | MRI (n=7) | MRI (n=4) | CT (n=2), MRI (n=7) | MRI (n=15) | CT (n=1) | CT (n=19), MRI (n=37) | |
| Electrode models | MDT 3387 (n=4), MDT 3389 (n=3) | MDT 3389 (n=11) | MDT 3387 (n=1), MDT 3389 (n=1) | MDT 3387 (n=7) | MDT 3387 (n=4) | MDT 3387 (n=2), MDT 3389 (n=6), MDT 3391 (n=1) | MDT 3387 (n=1), MDT 3389 (n=14) | MDT 3387 (n=1) | MDT 3387 (n=20), MDT 3389 (n =35), MDT 3391 (n=1) | |
| **Demographics** | |  |  |  |  |  |  |  |  |  |
| Sex (F/M) | 2F/5M | 4F/7M | 1F/1M | 1F/6M | 0F/4M | 3F/6M | 3F/12M | * | 15F/41M | |
| Age at surgery (mean ± SD, in years) | 31.4 ± 13.3 | N/A | 20.5 ± 9.2 | 27.7 ± 10.0 | 21.25 ± 2.1 | 28.2 ± 9.1 | 31.4 ± 9.9 | * | 29.2 ± 10.3 | |
| **Clinical symptom assessments** | |  |  |  |  |  |  |  |  |  |
| **Tic Score** | TTSS (/50) | YGTSS (/100) | YGTSS (/100) | YGTSS (/100) | YGTSS (/100) | YGTSS (/100) | YGTSS (/100) | YGTSS (/100) | TTSS (/50) | YGTSS (/100) |
| Baseline (mean ± SD) | 40.3 ± 5.3 | 63.4 ± 14.0 | 94.5 ± 0.7 | 87.3 ± 6.5 | 66.5 ± 14.0 | 84.2 ± 13.5 | 88.1 ± 9.1 | * | 40.3 ± 5.3 | 80.0 ± 15.2 |
| DBS ON (mean ± SD) | 14.4 ± 5.9 | 28.8 ± 20.0 | 43.0 ± 4.2 | 32.7 ± 27.0 | 41.8 ± 13.1 | 37.2 ± 20.1 | 58.7 ± 14.0 | * | 14.4 ± 5.9 | 42.7 ± 22.0 |
| Improvement (%) (mean ± SD) | 63.9 ± 14.2 | 55.2 ± 27.3 | 54.5 ± 4.2 | 62.9 ± 30.3 | 32.3 ± 32.4 | 53.8 ± 27.7 | 32.7 ± 17.7 | 0 ± 0 | 48.3 ± 26.6 | |
| **YBOCS** |  |  |  |  |  |  |  |  |  |  |
| Available Observations | n=2 | N/A | N/A | N/A | N/A | n=9 | n=11 | N/A | n=22 | |
| Baseline (mean ± SD) | 25.5 ± 7.8 |  |  |  |  | 31.6 ± 5.8 | 19.5 ± 6.9 |  | 25 ± 8.5 | |
| FU (mean ± SD) | 6.5 ± 9.2 |  |  |  |  | 21.2 ± 7.4 | 15.7 ± 7.5 |  | 17.1 ± 8.4 | |
| Improvement (%) | 67.5 ± 46.0 |  |  |  |  | 31.5 ± 22.2 | 22.0 ± 19.4 |  | 30.0 ± 25.2 | |

| **Target** | **STN** | | | | | **Full TS Cohort** | |
| --- | --- | --- | --- | --- | --- | --- | --- |
| **Cohort** | Cologne | Milan/Pisa | Shanghai | **STN Full Cohort** | |  |  |
| Surgical center | University Hospital Cologne | Fondazione IRCCS Instituto Neurologico Carlo Besta Milan | Ruijin Hospital Shanghai |  |  |  |  |
| Target | limbic part of STN, H2, VOP/VOA | STN | STN |  |  |  |  |
| Number of patients | 2 | 4 | 10 | 16 | | 115 | |
| Median Time at FU (months) | 6 | 6 | 6 | 6 [6-6] | | 6 [6-8] | |
| Related citation | ^1,5^ | ^7^ | ^8^ |  |  |  |  |
| **Imaging and electrode specifications** | |  |  |  |  |  |  |
| Postoperative imaging modality | CT (n=2) | CT (n=4) | CT (n=10) | CT (n=16) | | CT (n=70), MRI (n=45) | |
| Electrode models | MDT 3387 (n=1), MDT 3389 (n=1) | MDT 3389 (n=4) | MDT 3387 (n=2), SR 1210 (n=7), PINS L302 (n=1) | MDT 3387 (n=3), MDT 3389 (n=5) SR1210 (n=7), PINS L302 (n=1) | | MDT 3387 (n=44), MDT 3389 (n=57), MDT 3391 (n=1), NP DL-344-3.5 (n=5), SR1210 (n=7), PINS L302 (n=1) | |
| **Demographics** |  |  |  |  |  |  |  |
| Sex (F/M) | 0F/2M | 2F/2M | 1F/9M | 3F/13M | | 31F/84M | |
| Age at surgery (mean ± SD, in years) | 50 ± 17.0 | 33 ± 9.8 | 24.9 ± 10.4 | 30.1 ± 13.3 | | n=104, 29.5 ± 10.4 | |
| **Clinical symptom assessments** | |  |  |  |  |  |  |
| **Tic Score** | TTSS (/50) | YGTSS (/100) | YGTSS (/100) | TTSS (/50) | YGTSS (/100) | TTSS (/50) | YGTSS (/100) |
| Baseline (mean ± SD) | 36 ± 14.1 | 89.3 ± 5.3 | 69.7 ± 10.2 | 36 ± 14.1 | 75.3 ± 12.7 | 39.8 ± 7.5 | 78.6 ± 16.5 |
| DBS ON (mean ± SD) | 30 ± 9.9 | 34,25 | 25,7 | 30 ± 9.9 | 28.1 ± 17.1 | 20.5 ± 9.1 | 43.4 ± 23.4 |
| Improvement (%) (mean ± SD) | 15.6 ± 5.7 | 61.5 ± 17.8 | 62.9 ± 26.3 | 56.6 ± 27.1 | | 45.1 ± 26.6 | |
| **YBOCS** |  |  |  |  |  |  |  |
| Available Observations | N/A | N/A | N/A | N/A | | 38 | |
| Baseline (mean ± SD) |  |  |  |  |  | 22.0 ± 8.7 | |
| FU (mean ± SD) |  |  |  |  |  | 17.4 ± 8.1 | |
| Improvement (%) |  |  |  |  |  | 17.4 ± 35.3 | |

| **Cohort** | **Validation Cohort** |
| --- | --- |
| Surgical center | University Hospital Cologne |
| Target | CM/Voi |
| Number of patients | 8 |
| Median Time at FU (months) | 24.5 [11.0-64.3] |
| Related citation | - |
| **Imaging and electrode specifications** | |
| Postoperative imaging modality | CT (n=8) |
| Electrode models | MDT 3389 (n=3), MDT 3387 (n=1), MDT B33005 (n=1), MDT 33015 (n=3) |
| **Demographics** |  |
| Sex (F/M) | 1F/7M |
| Age at surgery (mean ± SD, in years) | 25.4 ± 3.6 |
| **Clinical symptom assessments** | |
| **Tic Score** | YGTSS (/100) |
| Baseline (mean ± SD) | 83.1 ± 12.3 |
| DBS ON (mean ± SD) | 35.4 ± 27.0 |
| Improvement (%) mean, std | 57.5 ± 31.6 |
| **YBOCS** |  |
| Available Observations | N/A |
| Baseline (mean ± SD) |  |
| FU (mean ± SD) |  |
| Improvement (%) |  |

*In cases where there is only one patient per center, the clinical and demographic information were not shared to avoid potential identifying details. Abbreviations: FU, Follow Up; TTSS, Total Tic Severity Scale (measures motor and phonic tic severity); YGTSS, Yale Global Tic Severity Scale^9^ (measures motor and phonic tic severity, as well as overall impairment); YBOCS, Yale-Brown Obsessive Compulsive Scale.^10^

#### **Modelling of the subcortical DBS white matter atlas**

#### **Anatomical Pathway Modelling**

To place the voxel-wise findings into an anatomical framework, it was required to first derive a precise anatomical understanding by developing a 3D streamlined model of the tracts in the pallidam/thalamic region. For this, we first compiled a list of subcortical pathways that could have potential relevance for GTS-DBS and reviewed anatomical and histological evidence from the human and non-human primate literature that described the trajectory of each tract of interest in relation to the surrounding anatomy (Supplementary Table 2, Supplementary Figs 2-9). In case of non-human primate studies, the homology across structures was determined based on the anatomical literature.^11–13^ Relevant anatomical landmarks were determined in standard template space with the help of atlases.

Connections of interest in GTS-DBS, such as CM-Pf efferents and pallidothalamic projections, remain largely underrepresented in the existing tractography literature, as it can be challenging to reconstruct using conventional tractography due to complex fiber architecture and the strong diffusion signal within the internal capsule which is orthogonally crossed by multiple of the thin candidate tracts of interest. To overcome this challenge, we systematically translated the tracts of interest into template space using manually curated trajectories^14^, building on an approach introduced by Petersen et al.^15^ This required to create a new software tool, CurveToBundle, which samples uniformly distributed streamlines along a trajectory, defined by curves in 3D space. In 3D Slicer, we first manually created curves in standard template space, following the trajectories described in anatomical literature. (Supplementary Figure 2C) By using *waypoints* with a defined spread value along the curve and interpolating between them, we defined the spatial boundaries of the bundle. Each tract was spatially translated from the centerline by a random amount following a uniform distribution inside the boundaries. Then, the tracts were displaced by sine curves, giving a less rigid appearance and more real-like looking streamlines. Finally, four types of anatomical constraints were applied when necessary: tracts must start from inside a specified nucleus; tracts must end inside a specified nucleus; tracts must go through specified nuclei; or tracts must avoid specified nuclei. (Supplementary Fig. 1D) Each tract was modelled in the left hemisphere and flipped to the right. The trajectories, spreads and the anatomical constraints based on anatomical literature are presented in detail in Supplementary Table 2 and Supplementary Figures 3-9.

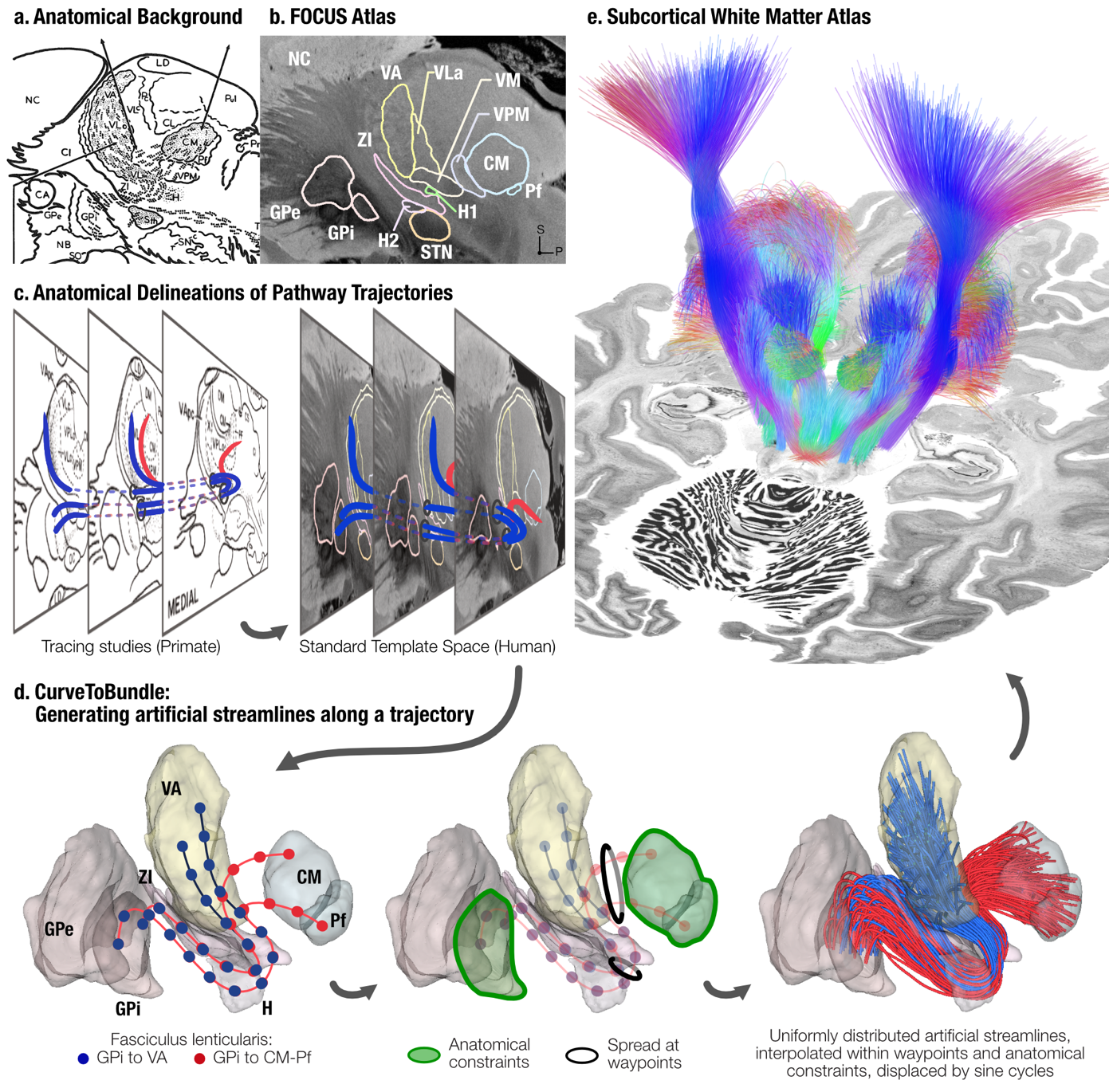
**Supplementary Figure 2: Delineation of a-priori tract candidates led to a 3D subcortical DBS atlas in stereotactic standard space.**

**a)** To trace anatomical connections of potential relevance for TS-DBS, we reviewed the anatomical literature and gathered numerous studies that depicted trajectories. The example image shows descriptions of ansa and fasciculus lenticulares in sagittal plane, adapted from ^16^. **b)** Sagittal view of the homologous anatomical structures shown in human standard template space. Relevant landmark structures were determined in standard template space with the help of a high quality novel subcortical atlas developed by our laboratory, which is described elsewhere^17^, as well as an ex-vivo human brain visualized by 7T-MRI.^18^ **c)** The trajectories of the pathways were translated to standard template space, amalgamating the descriptions from tracing studies in non-human primates (as depicted here), as well as evidence from dissections, histology and imaging studies in humans. In case of non-human primate studies, the homology across structures were determined based on the anatomical literature ^11–13^. Left panel adapted from Kuo and Carpenter, 1973 ^19^. Blue curves represent the trajectory of fasciculus lenticularis to ventral anterior thalamic nucleus (VA), red curves represent the collaterals of the same bundle to the CM-Pf complex. **d)** The trajectories were delineated as curves in 3D Slicer.^20^ Using a new tool developed for this study, CurveToBundle, streamline derivatives of this curve were sampled along the trajectory, adhering to anatomical limitations and dispersing into volumetric definitions of seed and endpoint nuclei. **e)** Multiple bundles were meticulously traced (see Supplementary Material), which together result in a subcortical white matter atlas that we make openly available as part of this work. Tracts were colored with respect to directionality (x-y-z direction mapping to r-g-b respectively) and are displayed on top of an axial slice from the BigBrain template ^21,22^. **Abbreviations.** NC, Nucleus Caudatus; GPe, Globus Pallidus Externa; GPi, Globus pallidus interna; ZI, Zona Incerta; H, H1, H2, Fields of Forel; VA, Ventral Anterior Nucleus; VLa, Ventrolateral Anterior Nucleus; VM, Ventromedial Nucleus; VPM, Ventral Posteromedial Nucleus; CM, Centromedian Nucleus; Pf, Parafascicular Nucleus; STN, Subthalamic Nucleus.

##### **Supplementary Table 2: Tract List**

| **Striatopallidofugal Connections** | | | | | | | | |
| --- | --- | --- | --- | --- | --- | --- | --- | --- |
| Connection | Abbreviation | Ref. | Corr. Figure | Anatomical Constraints | | | | Number of Streamlines |
|  |  |  |  | Start | End | Inside | Outside |  |
| Caudate – GPi | Cd_to_gpi_cau | ^23–27^ | S3 | Caudate^a^ | GPi^b^ | - | Putamen^a^ | 250 |
|  | Cd_to_gpi_ros | ^23–27^ | S3 | Caudate^a^ | GPi^b^ | - | Putamen^a^ | 250 |
| Putamen – Gpi | Pu_to_gpi_cau | ^16,23–27^ | S3 | Putamen^a^ | GPi^b^ | - | - | 250 |
|  | Pu_to_gpi_ros | ^16,23–27^ | S3 | Putamen^a^ | GPi^b^ | - | - | 250 |
| Caudate – SN | Cau_to_SN_cau | ^23–29^ | S3 | Caudate^a^ | SN^c^ | - | STN^b^, Putamen^a^ | 250 |
|  | Cau_to_SN_ros | ^23–29^ | S3 | Caudate^a^ | SN^c^ | - | STN^b^, Putamen^a^, AC^d^ | 250 |
| Putamen – SN | Pu_to_SN_cau | ^23–29^ | S3 | Putamen^a^ | SN^c^ | - | STN^b^ | 250 |
|  | Pu_SN_ros | ^23–29^ | S3 | Putamen^a^ | SN^c^ | - | STN^b^, AC^d^ | 250 |
| Ventral Striatum – SN/VTA | venStr_to_snvta_overAC | ^23,24,28–32^ | S3 | Caudate^a^ | - | - | STN^b^, AC^d^, lateral SN | 125 |
|  | venStr_to_snvta_underAC | ^23,24,28–32^ | S3 | NAcc^a^ | - | - | STN^b^, lateral SN | 125 |
| **Efferents of CM-Pf Complex** | | | | | | | | |
| CM to dorsolateral postcomissural putamen | CM_to_Put | ^33–38^ | S4 | CM^c^ | Striatum^c^ | - | - | 500 |
| CM to STN | CM_STN | ^34,35,37^ | S4 | CM^c^ | STN^c^ | - | Associative zone of STN^b^ | 250 |
| Pf to Caudate Head | Pf_to_Cau_head | ^35–38^ | S5 | Pf^c^ | Caudate^a^ | - | Ventricles | 250 |
| Pf to Caudate Body and Tail | Pf_to_Cau_bodytail | ^35–38^ | S5 | Pf^c^ | Caudate^a^ | - | Ventricles, AC^c^ | 250 |
| Pf to Putamen | Pf_to_Put | ^35–38^ | S5 | Pf^c^ | Striatum^c^ | - | STN^c^, AC^c^ | 250 |
| Pf to Ventral Striatum | Pf_to_venStr | ^35–38^ | S5 | Pf^c^ | Striatum^c^ | - | STN^c^, AC^c^ | 250 |
| Pf to Pallidum | Pf_to_Pall | ^35–38^ | S5 | Pf^c^ | - | - | STN^c^ | 50 |
| Pf to STN | Pf_to_STN | ^35,37^ | S5 | Pf^c^ | STN^c^ | - | - | 250 |
| Pf to VTA | Pf_to_VTA | ^35,37^ | S5 | Pf^c^ | - | - | RN^c^, STN^c^ | 250 |
| **Pallidosubthalamic Connections** | | | | | | | | |
| Ansa Subthalamica | Ansa_subthalamica | ^16,39,40^ | S6 | STN^b^ | GPi^b^ | - | - | 250 |
| GPi – STN | GPi_stn | ^15,41^ | - | GPi^c^ | STN^c^ | - | - | 250 |
| GPe – STN | GPe_stn | ^15,41,42^ | - | GPe^c^ | STN^c^ | - | - | 250 |
| **Pallidothalamic Connections** | | | | | | | | |
| Ansa Lenticularis to VA | AL_to_va | ^13,16,19,23,41,43–50^ | S6-8 | GPi^c^ | VA^c^ + VLa^c^ | AL^c^ | STN^c^, VLp^c^ | 250 |
| Ansa Lenticularis to CM-Pf | AL_to_cmpf | ^16,19,23,41,43,45–49^ | S8 | GPi^c^ | CM^c^ + Pf^c^ | AL^c^ | STN^c^ | 250 |
| Fasciculus Lenticularis to VA | FL_to_va | ^16,41,43,19,47,49,50^ | S2, S6-8 | GPi^c^ | VA^c^ + VLa^c^ | - | STN^c^, VLp^c^ | 250 |
| Fasciculus Lenticularis to CM-Pf | FL_to_cmpf | ^16,19,41,43,47,49^ | S2, S8 | GPi^c^ | CM^c^ + Pf^c^ | H2^c^ | STN^c^ | 250 |
| **Cerebellothalamic Connections** | | | | | | | | |
| dDRTT – dorsal dentate to VL, M1 projecting zones | dDRTT_VL_ros | ^23,44,48,13,49–53^ | S6,7, 9 | DN^c^ | VLp^c^ | - | VPL^c^, VPM^c^, ventral RN | 250 |
| dDRTT – dorsal dentate to VL, SMA projecting zones | dDRTT_VL_cau | ^23,44,48,13,49–54^ | S6,7,9 | DN^c^ | VLp^c^ | - | VPL^c^, VPM^c^, ventral RN | 250 |
| dDRTT – ventral dentate to VL/MD | dDRTT_ventral | ^23,44,48,50–53,55–57^ | S6,7,9 | DN^c^ | Thal^c^ | - | CM^c^, VPM^c^, ventral RN | 250 |
| **Sensory Afferents** | | | | | | | | |
| Medial Lemniscus | Medial_lemn | ^23,44,58–60^ | S6 | VPL^c^ | - | - | - | 250 |
| Trigeminal Lemniscus | Trigeminal_lemn | ^23,44,58–60^ | S6 | VPM^c^ | - | - | - | 250 |
| **Corticospinal Tract** | | | | | | | | |
| CST | CST | ^15,23,61,62^ | S2 | BA4^e^ | - | - | GPi^b^, STN^b^ | 1000 |

**Atlases:** ^a^ - CIT168 Reinforcement Learning Atlas ^63^, ^b^ - DISTAL atlas^64^, ^c^ - FOCUS atlas^17^, ^d^ - Atlas of the Human Hypothalamus^65^, ^e^ - Digitized Brodmann Atlas^66^ **Abbreviations:** Ref, References; ros, rostral; cau, caudal; AC, anterior commissure; AL, ansa lenticularis; BA4, Brodmann Area 4; CM, Centromedian nucleus; dDRTT, decussating dentro-rubro-thalamic tract; DN, dentate nucleus; GPe, external pallidum; GPi, internal pallidum; H2, Fields of Forel H2; M1, primary motor cortex; MD, mediodorsal nucleus; NAcc, nucleus accumbens; Pf, Parafascicular nucleus; RN, red nucleus; SMA, supplementary motor area; SN, Substantia Nigra; SNr, Substantia Nigra pars reticularis; STN, Subthalamic Nucleus; Thal, thalamus; VA, ventral anterior thalamic nucleus; VL, ventral lateral thalamic nucleus; VLa, ventral lateral nucleus anterior portion; VLp, Ventral lateral nucleus posterior portion; VPL, ventral posterolateral nucleus; VPM, ventral posteromedial nucleus; VTA, ventral tegmental area

##### Example tract modelling steps

For centromedian nucleus (CM) projections to dorsolateral post commissural putamen, the trajectory and the endpoints of the bundle, as demonstrated by the anterograde tracer injections into CM by Sadikot and colleagues^35,37^, were segmented in standard template space. Two curves that leave CM rostrally and continue through ventral reticular thalamic nucleus were generated. Both curves were, then, continued within internal capsule. The first curve defining the rostral boundary ends in rostral portion and the second one ends in caudal portion of the segmented area, which corresponds to dorsolateral post-commissural putamen. 500 streamlines were generated in a uniform distribution in between these two curves. Spread values were interpolated with a 3^rd^ order spline, with the range outside the *waypoints* being extrapolated. Streamlines were displaced by 1.5 sine cycles. CM from Focus atlas was set as a starting point constraint and Striatum from Focus atlas was set as an endpoint constraint. Uniformly generated streamlines were trimmed according to these anatomical constraints. These steps were then repeated for each potential fiber bundle, resulting in a comprehensive subcortical white matter atlas.

##### Anatomical Pathway Descriptions

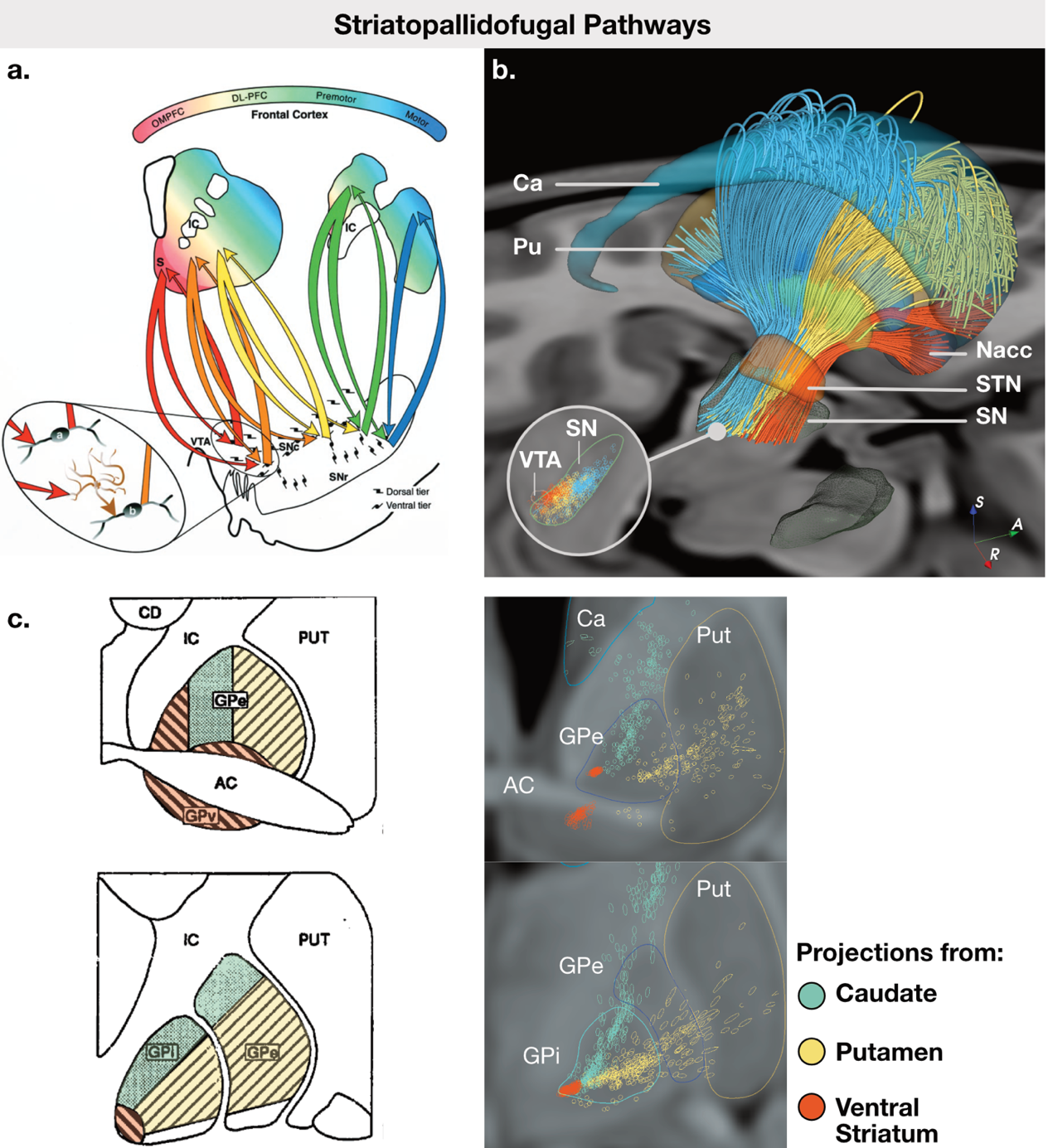

**Supplementary Figure 3: A)** Striatopallidofugal fibers are organized in a rostrocaudal gradient at the striatal level, according to the cortical input areas. In the subcortical level, the efferents take a mediolateral functional organization. Figure adapted from Haber et al., 2000.^28^ **B)** The rostrocaudal to mediolateral gradient is reflected in our tract models. Colors represent the functional gradient: red, limbic zones; yellow, associative zones; blue, motor zones. **C)** In addition to the rostrocaudal to mediolateral organization, the fibers are organized according to the input location at the pallidal level. Fibers originating from caudate nucleus traverse on dorsal regions (green), fibers from putamen traverse ventrally (yellow) and ventral striatal efferents traverse in areas marked by red. It should be noted that colors represent fiber origin here. Figure adapted from Hazrati and Parent, 1992.^24^ (Left panel) This organization was reflected in our fiber model. (Right panel) 3D models are taken from DISTAL^64^ and CIT168 reinforcement learning^63^ atlases.

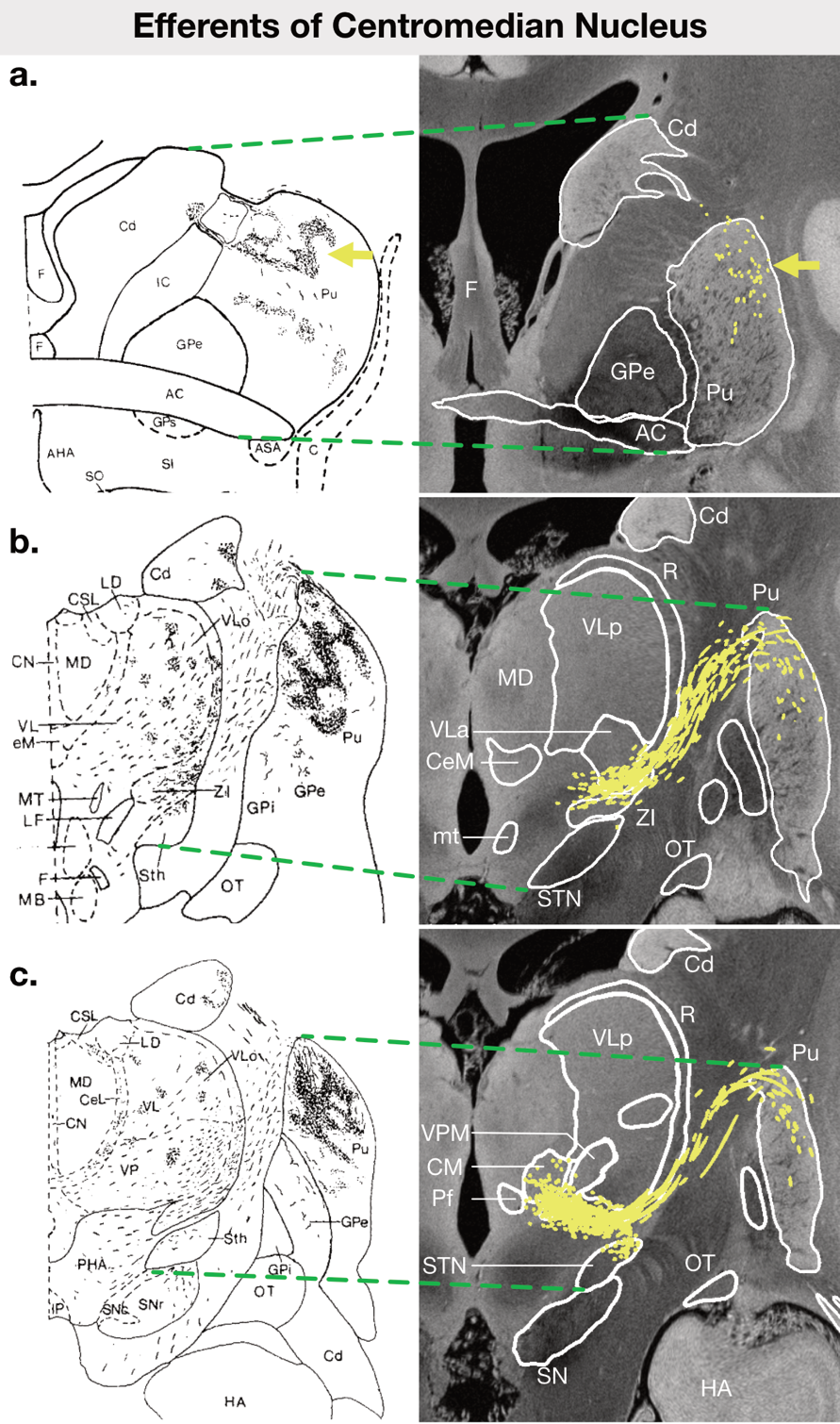

**Supplementary Figure 4: Efferents of Centromedian Nucleus (CM)**. CM projects to dorsolateral post-commissural putamen (Pu, Panel A-C, yellow arrow) and caudal two thirds of subthalamic nucleus (STN, Panel C). Left column shows results of anterograde tracings from CM, reproduced with permission from Sadikot et al.^37^ Right column shows the corresponding coronal sections in humans in standard template space. The green dashed lines aim to guide the viewer’s eye to the corresponding anatomical structures. The backdrop is 7T post-mortem ultra-high resolution MRI image.^18^ The atlas structures outlined by white lines are from FOCUS atlas. The yellow lines show the trajectory of the modelled pathway in corresponding slices.

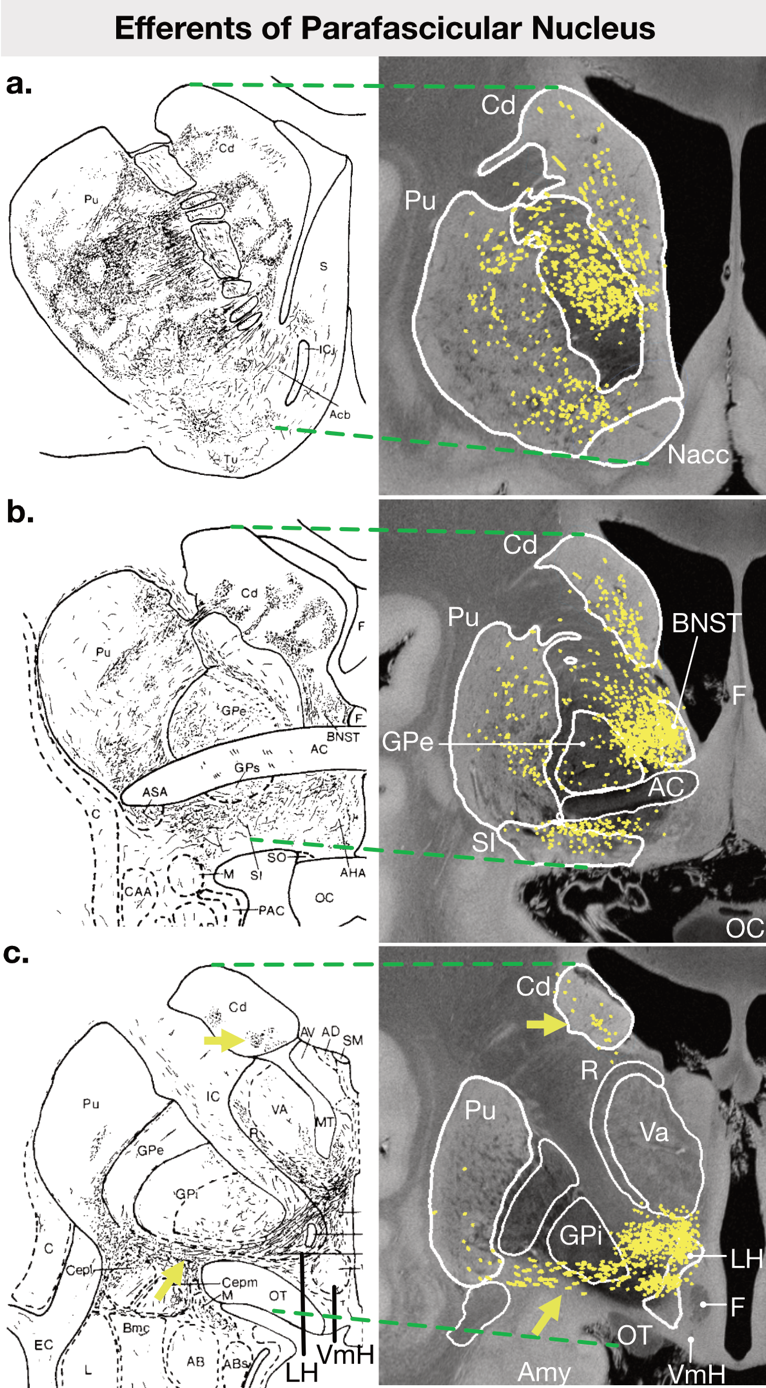

**Supplementary Figure 5: Efferents of Parafascicular Nucleus**

Parafascicular nucleus (Pf) projects to caudate nucleus (Cd), rostral putamen (Pu) and Nucleus Accumbens (Nacc), corresponding to associative-limbic zones of striatum.^23,35,67^ (Panel A-C) Projections travel through bed nucleus of stria terminalis (BNST) and lateral hypothalamic region (LH) and extend collaterals to external pallidum (GPe, Panel B,C) and internal pallidum (GPi, Panel C, bottom yellow arrow). Left column shows results of anterograde tracings from Pf, reproduced with permission from Sadikot et al.^37^ Right column shows the corresponding coronal sections in humans in standard template space. Green dashed lines aim to guide the viewer’s eye to the corresponding anatomical structures. The backdrop is 7T post-mortem ultra-high resolution MRI image.^18^ The atlas structures outlined by white lines are from FOCUS atlas. The yellow lines show the trajectory of the modelled pathway in corresponding slices.

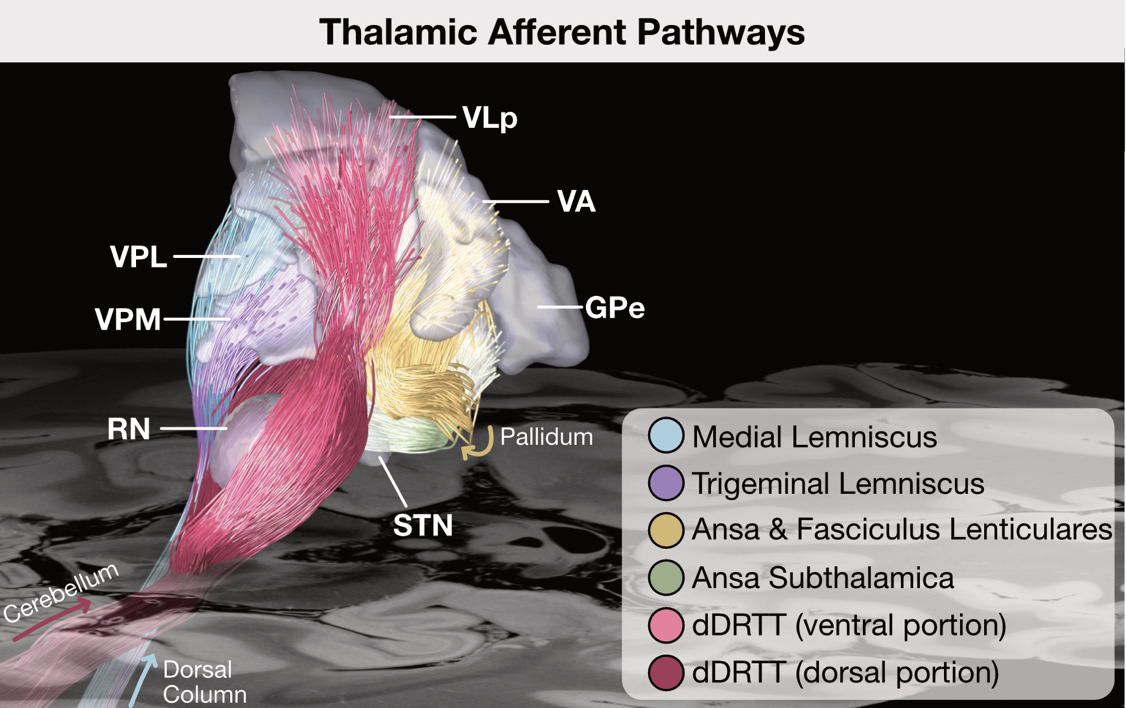

**Supplementary Figure 6: Organization of Thalamic Afferents**

Thalamic input pathways are displayed in sagittal 3D view. Pallidal input (ansa and fasciculus lenticulares) traverses from fields of Forel and enters ventral anterior nucleus (VA). Cerebellar input from contralateral dentate nucleus wraps around red nucleus (RN) and enters into ventral lateral nucleus, posterior portion (VLp). Carrying sensory input from dorsal column nuclei, medial and trigeminal lemnisci enter into ventral posterolateral (VPL) and ventral posteromedial (VPM) nuclei respectively. The backdrop is 7T post-mortem ultra-high resolution MRI image.^18^

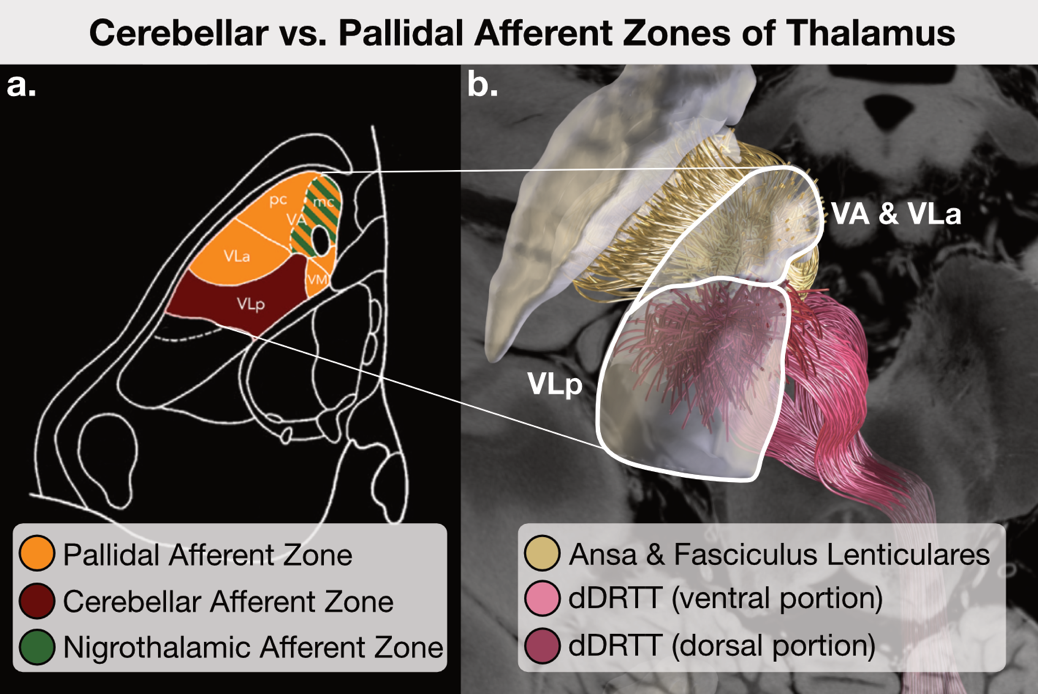

**Supplementary Figure 7: A)** Ventral anterior (VA), ventralis lateralis anterior portion (VLa) and Ventromedial (VM) nuclei form the pallidal afferent zones of thalamus. Ventralis lateralis posterior portion (VLp) is the cerebellar afferent portion. Figure reproduced with permission from Neudorfer et al.^13^ **B)** Our manually created pathways reflect this principle of organization, displayed in coronal 3D view.

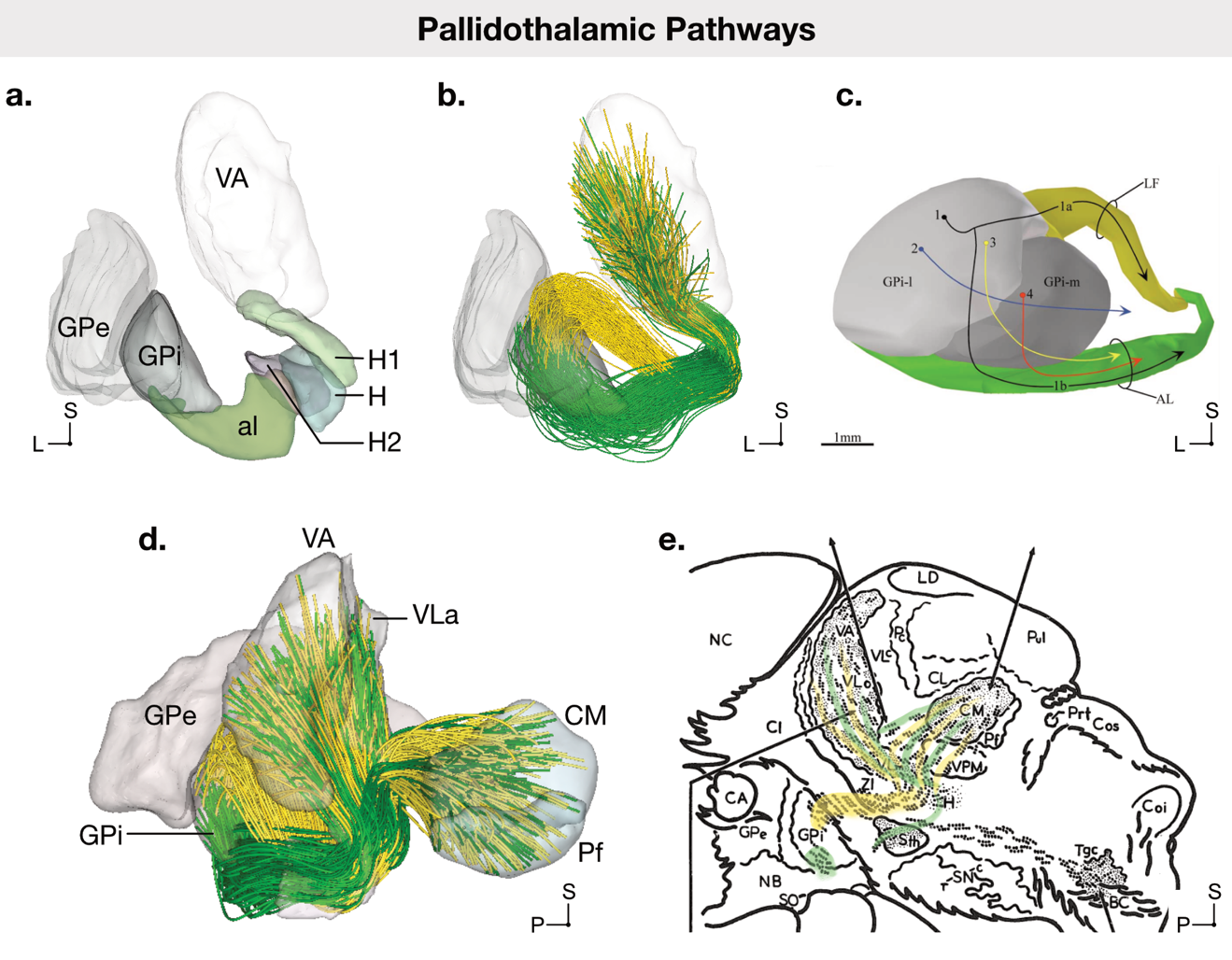

**Supplementary Figure 8: A)** 3D representation of histology-based segmentations of ansa lenticularis (al) and fields of Forel (H, H1, H2). **B)** Pallidothalamic pathways modeled in 3D space. Ansa lenticularis (green) exits internal pallidum (GPi) on its ventral border, travels through H and H1 to enter ventral anterior (VA) and ventral anterior lateralis (VLa) nuclei of thalamus. Fasciculus lenticularis (yellow) leaves internal pallidum (GPi) on its dorsal border, traverses through H2, corresponding to the dorsal region of STN, and joins ansa lenticularis at Field of Forel H. **C)** Single-axon studies in monkeys have demonstrated the dorsal and ventral respective trajectories,^45^ in line with previous evidence from non-human primates^16,43,46^ and humans.^41^ Note the resemblance of this anatomical description to our sweetspot models (Figure 2B, peaks P1 and P2). Figure adapted with permission from ^45^. **D)** Our models of pallidothalamic fibers. After merging at H, pallidothalamic bundles pass through Ventromedial nucleus (VM), reaching to VA and VLa. The collaterals to CM-Pf complex extend in the sagittal plane at the level of VM, traversing above and through VPM. E) Our models follow the literature descriptions as closely as possible. Figure adapted with permission from ^16^. Note the resemblance of the trajectory described in (D-E) with the thalamic tic sweetspot peaks (Figure 2B, peaks T1, T2).

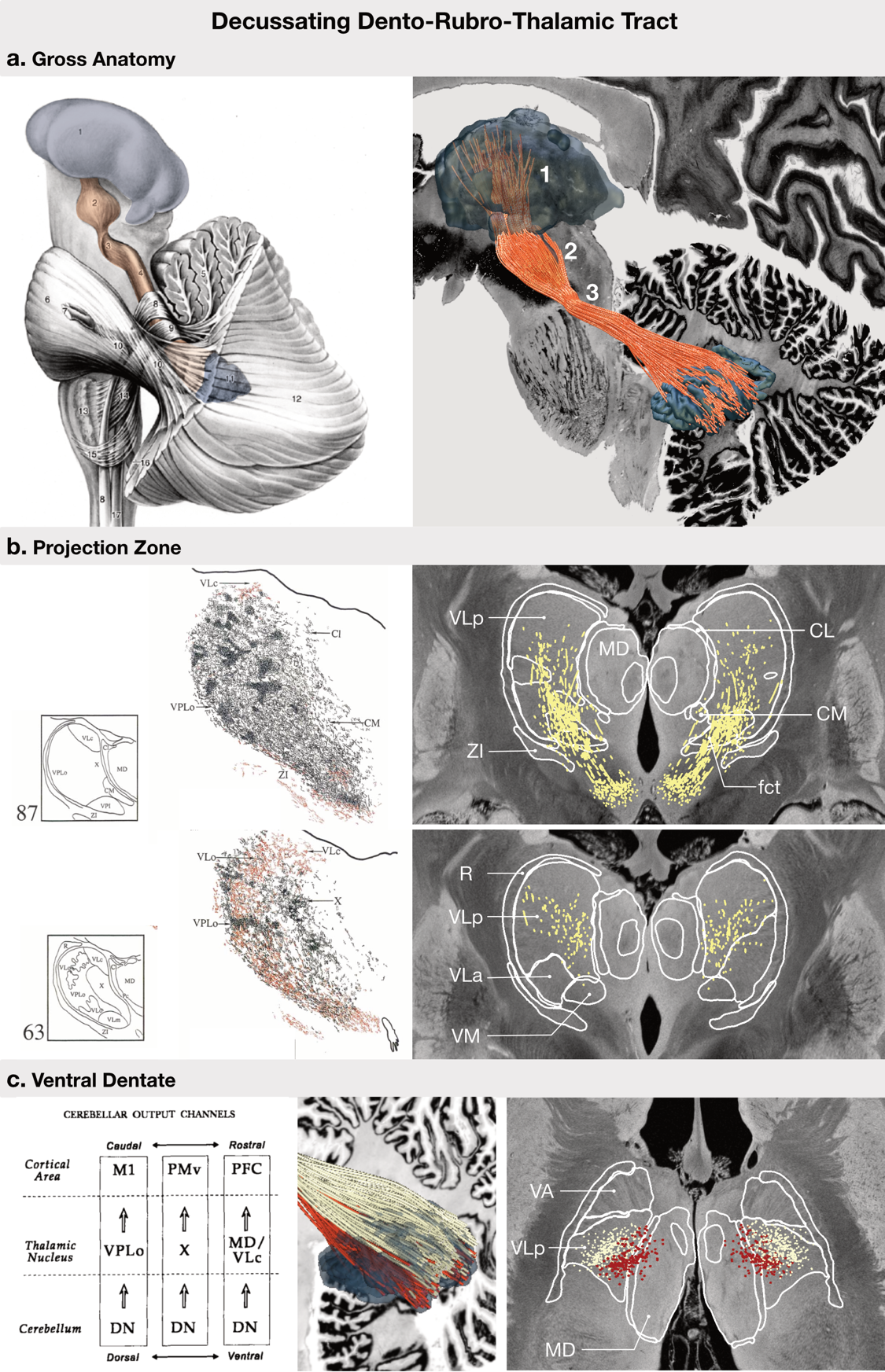
**Supplementary Figure 9: Cerebellothalamic Pathways**

**A)** Decussation dento-rubro-thalamic tract (dDRTT, orange) originates from dentate nucleus, leaves cerebellum in superior cerebellar peduncle, crosses to the contralateral hemisphere (3), wraps around red nucleus (2) and enters thalamus (1). Left panel reproduced from Nieuwenhuys et al.^23^ Right panel shows the gross anatomy followed by our dDRTT model (orange), displayed on top of a sagittal slice from BigBrain template^21,22^. **B)** Left panel shows coronal sections after anterograde injections into dentate nucleus (black) and pallidum (red) in non-human primate models, reproduced with permission from ^48^. VPLo and VLc structures from non-human primate terminology corresponds to ventral and dorsal parts of ventral lateral nucleus posterior portion (VLp), respectively^11–13^. Right panel shows the entry of dDRTT models into thalamus via cerebellothalamic fascicle (fct) and innervation of VLp on coronal panels overlaid on ^18^. Note that pallidal regions, namely VLo corresponding to ventral lateral anterior (VLa) in humans, are not covered by dDRTT models. **C)** dDRTT shows a ventrodorsal functional organization.^51,53^ Left panel, reproduced from ^51^, illustrates that dorsal dentate nucleus (DN) synapses in regions of thalamus that project to primary motor cortex (M1), whereas ventral regions feed areas that project to prefrontal cortex (PFC). Middle panel shows our models of dorsal (white) and ventral (red) dDRTT in sagittal view. Right panel shows a coronal section illustrating the ventral dDRTT (red) extending into mediodorsal nucleus at dorsal levels. Of note, to the best of our knowledge, the functional gradient around red nucleus is not well explored in the literature. Therefore, in the red nucleus region, we adhered to the descriptions of ^23^ and modelled the ventral part to pass through lateral and rostral regions, and dorsal part to traverse near fasciculus retroflexus, at the dorsomedial portion of red nucleus.

### Supplementary Results

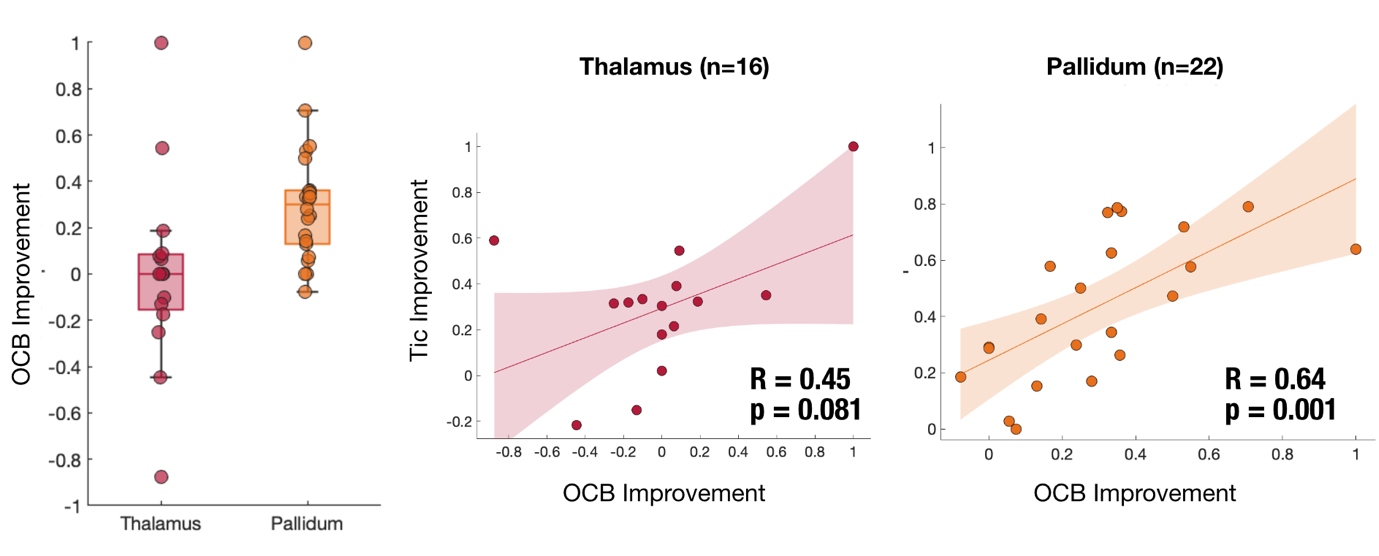

**Supplementary Figure 10: Obsessive-compulsive behavior improvement.** Pallidum cohort showed significantly more improvement in OCB domain compared to the thalamic cohort. (left panel) OCB improvement was significantly positively correlated in the pallidal cohort. (right panel) The association was not significant in the thalamic cohort. (middle panel)

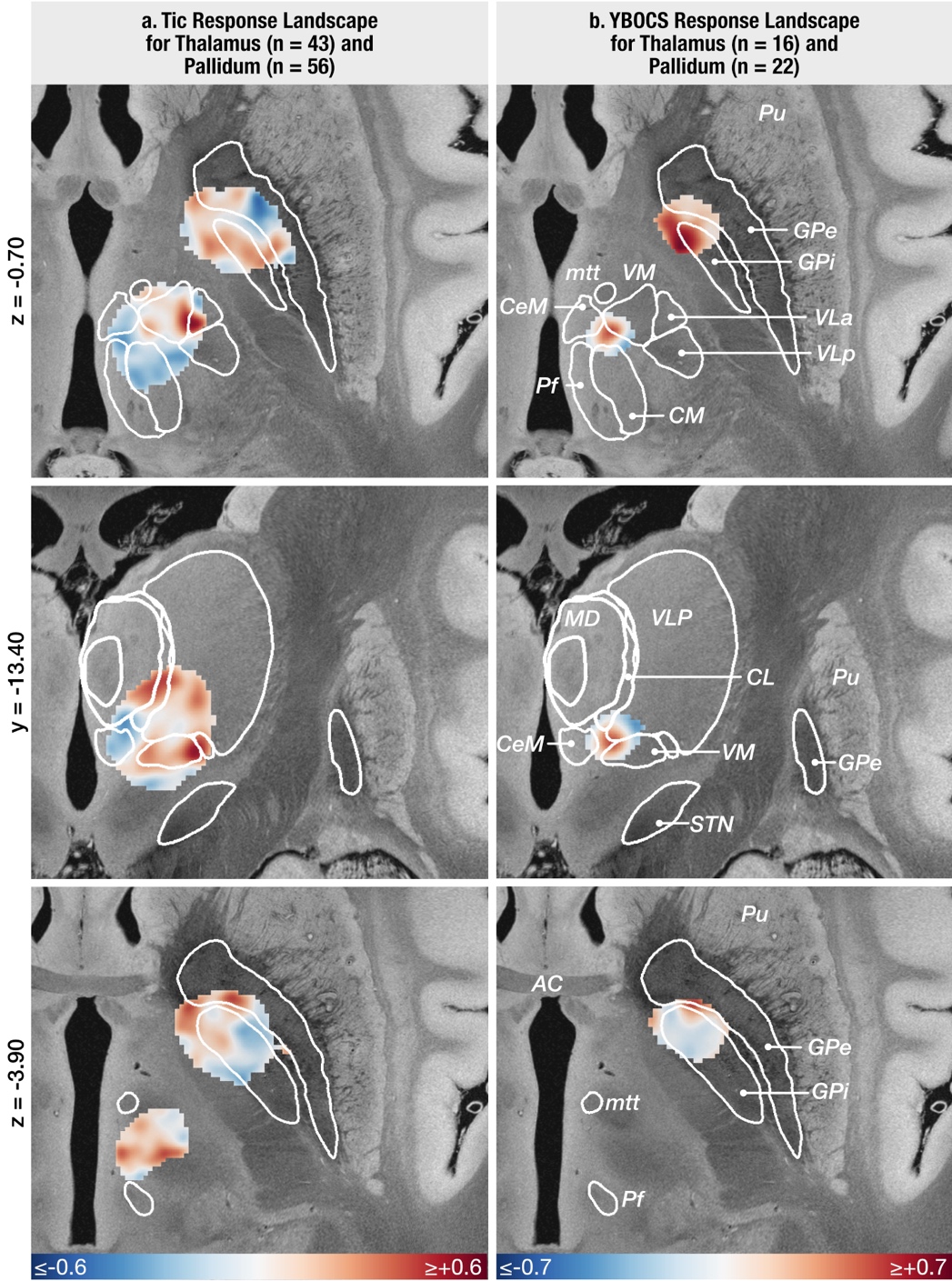

**Supplementary Figure 11: Tic vs. OCB response maps. A)** Tic response landscape of thalamus and pallidum, as equally shown in Figure 1 of the main text. **B)** YBOCS response landscapes of the same cohorts, shown in comparison to the tic response landscapes. The YBOCS response peak for pallidum overlapped with tic response peaks and extended slightly more anteriorly in dorsal sections, however, the YBOCS response peak for the thalamus anatomically diverged from the main tic response peaks.

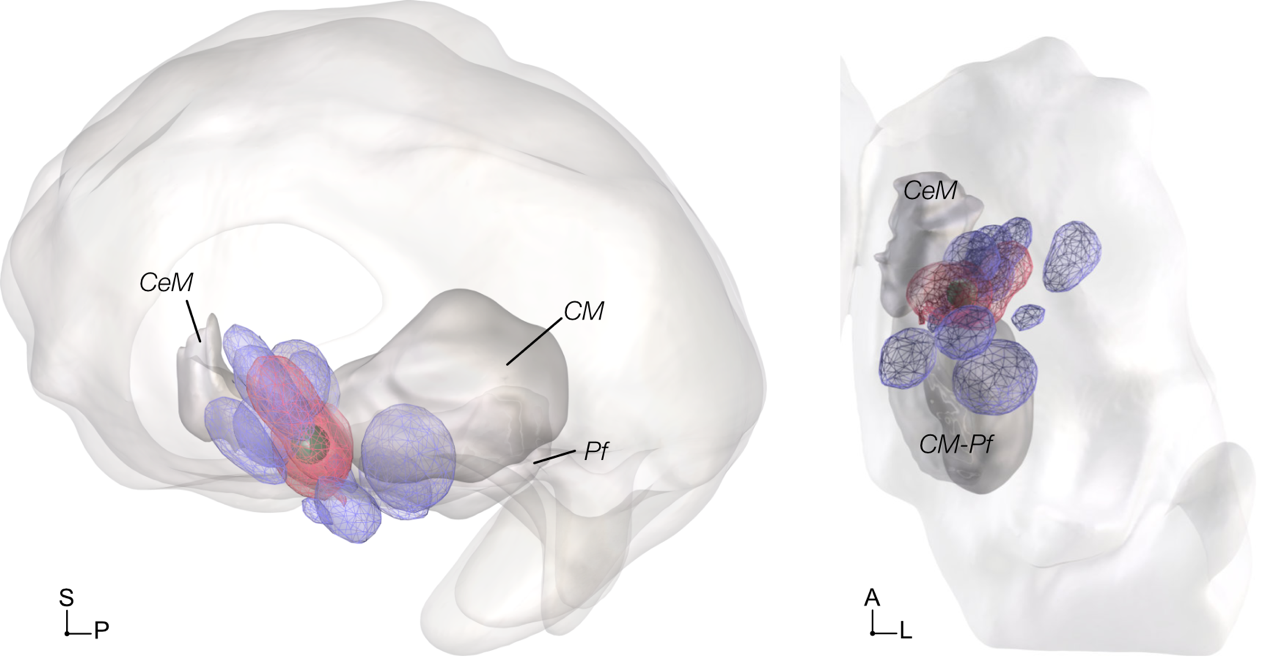

**Supplementary Figure 12: Top vs. Bottom OCB Responders from the thalamus cohort (n=16).** VTAs of top (pink) vs bottom (blue) responders from the thalamic cohort are displayed in sagittal (left) and axial (right) views. The stimulation volumes of three thalamic responders (YBOCS improvement >15%) form the thalamic OCB response peak (green sphere).

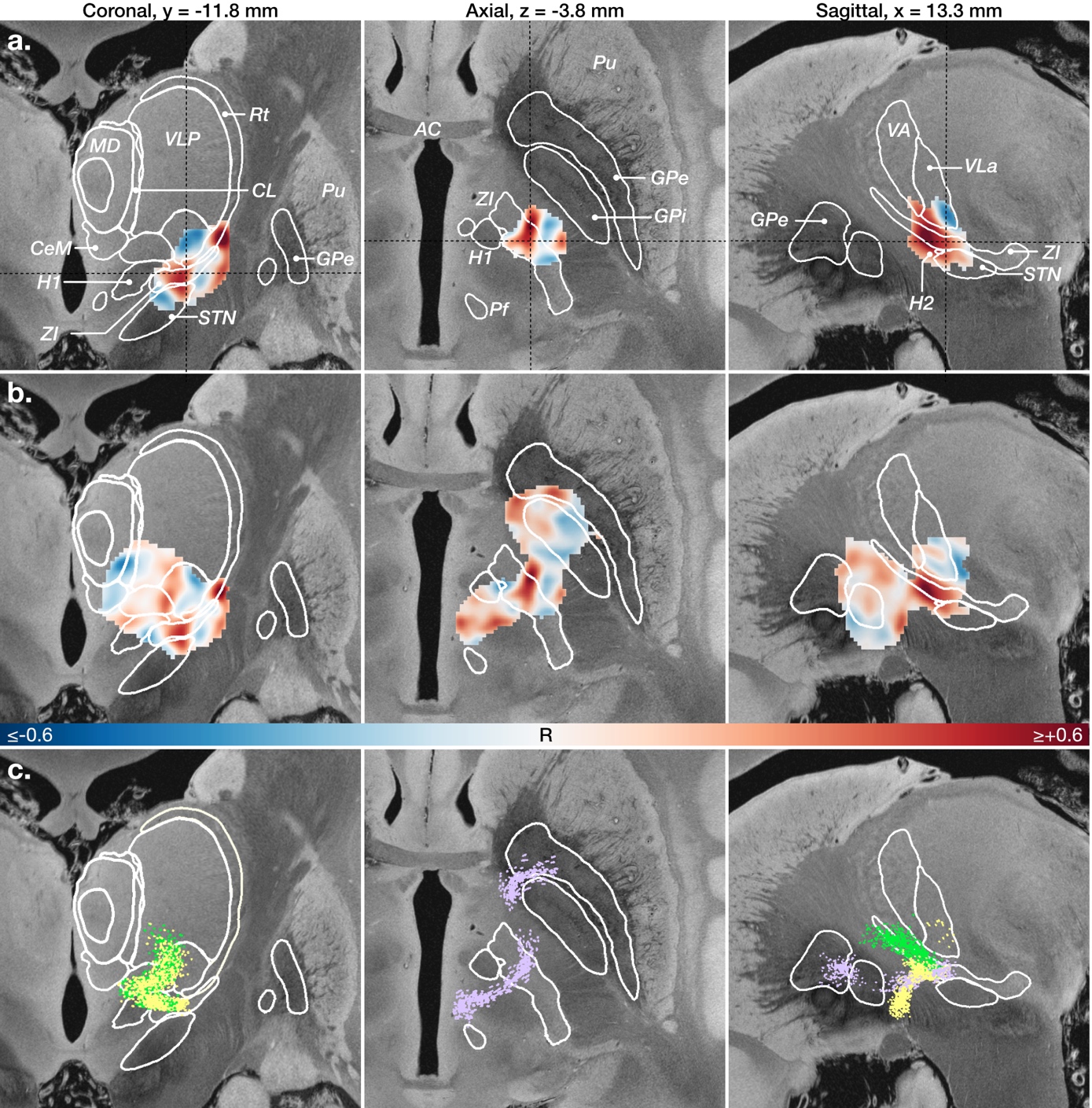

**Supplementary Figure 12: Tic response peaks of STN. a)** coronal, axial and sagittal views of the tic response map of STN cohort alone. (n=16) **b)** The E-fields of the full cohort formed a gradient, covering the area completely between thalamus, STN and pallidum. We therefore created a combined tic response map of the full cohort. (n=115) The addition of STN revealed two additional peaks: one in dorsal STN, extending mainly into the pathways of passage above STN and into zona incerta; another in reticular nucleus of thalamus. **c)** Candidate tic response pathways generated by tracing the voxel-like tic response peaks, adhering to anatomical descriptions. Pathways described here are ansa lenticularis (yellow), fasiculus lenticularis (green) and efferent pathways of intralaminar thalamic nuclei (purple).

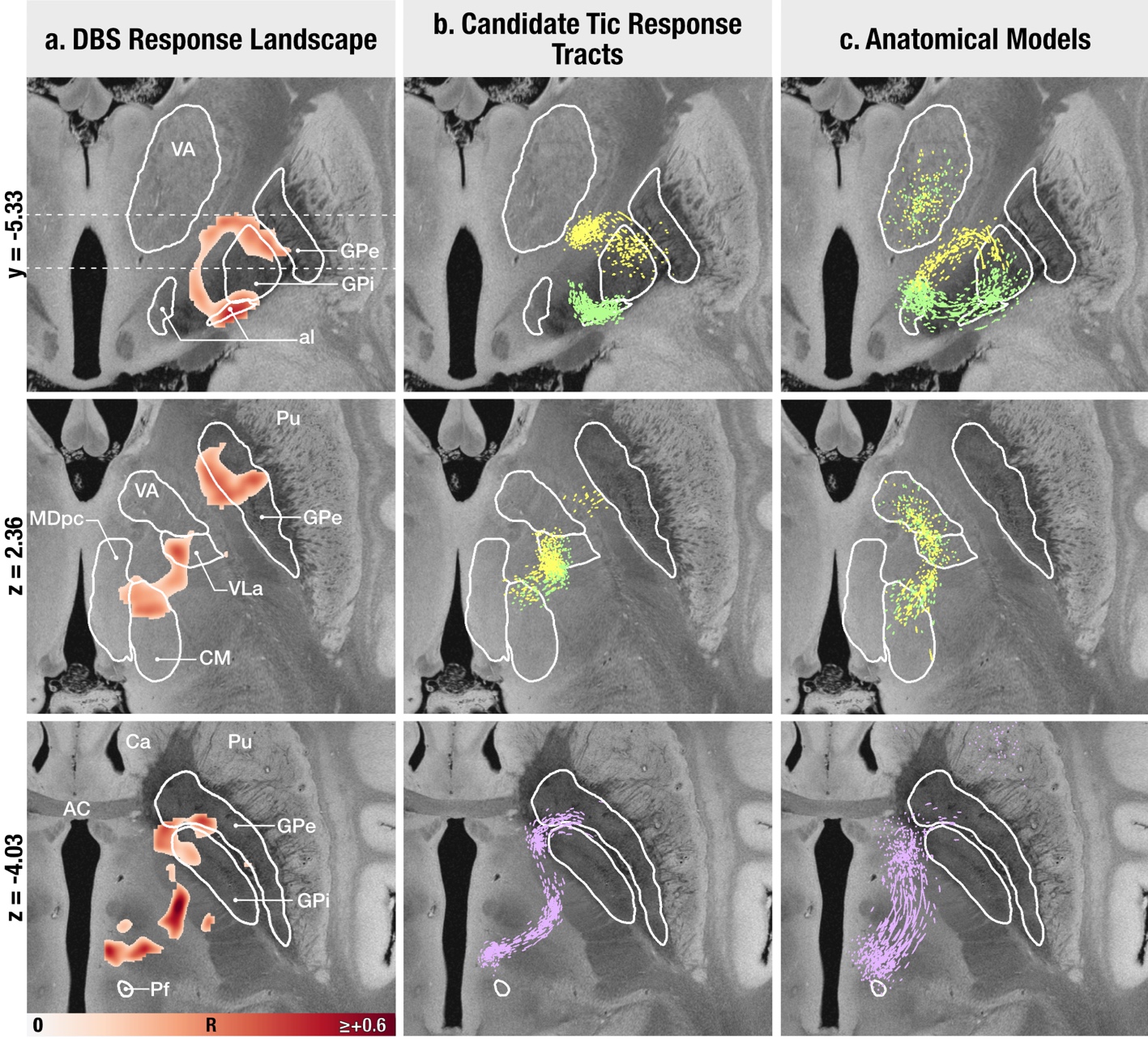

**Supplementary Figure 13: Sweetspot data and a-priori anatomical tracts.** Tract-like clusters in DBS response landscape (a) were retraced to create candidate tic response tracts (b) using CurveToBundle, bearing in mind the a-priori defined anatomical models (c). Yellow represents fasciculus lenticularis, green represents ansa lenticularis, purple represents Pf outflow bundle. Anatomical atlas structures are overlaid on 7T postmortem MRI image.^18^ Differences between the a-priori tracts (c) that were purely informed by the anatomical literature and the retraced tracts (b) which were mainly informed by the sweet spot landscape can be seen. A critical note on this matter is, however, that especially for ansa lenticularis and fascicularis, textbook knowledge specifically focuses on their motor domains (and tracer injections have often focused on posterior pallidal regions). Based on in-depth conversations with expert anatomists (M.P. & A.S.) who had carried out macaque tracer experiments in these regions, the trajectories of these bundles in reality are much more diffuse than can be appreciated in most textbooks (which typically focus on the most prominent parts of the structures that correspond to the motor domains).

### References

1.	Ganos C, Al-Fatly B, Fischer JF, et al. A neural network for tics: insights from causal brain lesions and deep brain stimulation. *Brain*. Published online January 13, 2022:awac009. doi:10.1093/brain/awac009

2.	Hollunder B, Ostrem JL, Sahin IA, et al. Mapping dysfunctional circuits in the frontal cortex using deep brain stimulation. *Nat Neurosci*. Published online February 22, 2024:1-14. doi:10.1038/s41593-024-01570-1

3.	Vilela-Filho O, Souza JT, Ragazzo PC, et al. Bilateral Globus Pallidus Externus Deep Brain Stimulation for the Treatment of Refractory Tourette Syndrome: An Open Clinical Trial. *Neuromodulation*. 2023;0(0). doi:10.1016/j.neurom.2023.04.473

4.	Johnson KA, Duffley G, Anderson DN, et al. Structural connectivity predicts clinical outcomes of deep brain stimulation for Tourette syndrome. *Brain*. 2020;143(8):2607-2623. doi:10.1093/brain/awaa188

5.	Baldermann JC, Hennen C, Schüller T, et al. Normative Functional Connectivity of Thalamic Stimulation for Reducing Tic Severity in Tourette Syndrome. *Biological Psychiatry: Cognitive Neuroscience and Neuroimaging*. 2022;7(8):841-844. doi:10.1016/j.bpsc.2022.01.009

6.	Johnson KA, Fletcher PT, Servello D, et al. Image-based analysis and long-term clinical outcomes of deep brain stimulation for Tourette syndrome: a multisite study. *J Neurol Neurosurg Psychiatry*. 2019;90(10):1078-1090. doi:10.1136/jnnp-2019-320379

7.	Vissani M, Cordella R, Micera S, Eleopra R, Romito LM, Mazzoni A. Spatio-temporal structure of single neuron subthalamic activity identifies DBS target for anesthetized Tourette syndrome patients. *J Neural Eng*. 2019;16(6):066011. doi:10.1088/1741-2552/ab37b4

8.	Dai L, Xu W, Song Y, et al. Subthalamic deep brain stimulation for refractory Gilles de la Tourette’s syndrome: clinical outcome and functional connectivity. *J Neurol*. 2022;269(11):6116-6126. doi:10.1007/s00415-022-11266-w

9.	Leckman JF, Riddle MA, Hardin MT, et al. The Yale Global Tic Severity Scale: Initial Testing of a Clinician-Rated Scale of Tic Severity. *Journal of the American Academy of Child & Adolescent Psychiatry*. 1989;28(4):566-573. doi:10.1097/00004583-198907000-00015

10.	Goodman WK, Price LH, Rasmussen SA, et al. The Yale-Brown Obsessive Compulsive Scale: I. Development, Use, and Reliability. *Arch Gen Psychiatry*. 1989;46(11):1006-1011. doi:10.1001/archpsyc.1989.01810110048007

11.	Ilinsky IA, Kultas-Ilinsky K. Sagittal cytoarchitectonic maps of the Macaca mulatta thalamus with a revised nomenclature of the motor-related nuclei validated by observations on their connectivity. *J Comp Neurol*. 1987;262(3):331-364. doi:10.1002/cne.902620303

12.	Barbas H, García-Cabezas MÁ, Zikopoulos B. Frontal-thalamic circuits associated with language. *Brain and Language*. 2013;126(1):49-61. doi:10.1016/j.bandl.2012.10.001

13.	Neudorfer C, Kultas-Ilinsky K, Ilinsky I, et al. The role of the motor thalamus in deep brain stimulation for essential tremor. *Neurotherapeutics*. 2024;21(3):e00313. doi:10.1016/j.neurot.2023.e00313

14.	Petersen MV, McIntyre CC. Comparison of Anatomical Pathway Models with Tractography Estimates of the Pallidothalamic, Cerebellothalamic, and Corticospinal Tracts. *Brain Connect*. 2023;13(4):237-246. doi:10.1089/brain.2022.0068

15.	Petersen MV, Mlakar J, Haber SN, et al. Holographic Reconstruction of Axonal Pathways in the Human Brain. *Neuron*. 2019;104(6):1056-1064.e3. doi:10.1016/j.neuron.2019.09.030

16.	Nauta WJH, Mehler WR. Projections of the lentiform nucleus in the monkey. *Brain Research*. 1966;1(1):3-42. doi:10.1016/0006-8993(66)90103-X

17.	Friedrich H, Sahin IA, Rajamani N, et al. A precise atlas of the human subcortex. *bioRxiv*. Preprint posted online February 15, 2026:2026.02.13.705755. doi:10.64898/2026.02.13.705755

18.	Edlow BL, Mareyam A, Horn A, et al. 7 Tesla MRI of the ex vivo human brain at 100 micron resolution. *Sci Data*. 2019;6(1):244. doi:10.1038/s41597-019-0254-8

19.	Kuo JS, Carpenter MB. Organization of pallidothalamic projections in the rhesus monkey. *Journal of Comparative Neurology*. 1973;151(3):201-235. doi:10.1002/cne.901510302

20.	Kikinis R, Pieper SD, Vosburgh KG. 3D Slicer: A Platform for Subject-Specific Image Analysis, Visualization, and Clinical Support. In: Jolesz FA, ed. *Intraoperative Imaging and Image-Guided Therapy*. Springer; 2014:277-289. doi:10.1007/978-1-4614-7657-3_19

21.	Amunts K, Lepage C, Borgeat L, et al. BigBrain: An Ultrahigh-Resolution 3D Human Brain Model. *Science*. 2013;340(6139):1472-1475. doi:10.1126/science.1235381

22.	Xiao Y, Lau JC, Anderson T, et al. An accurate registration of the BigBrain dataset with the MNI PD25 and ICBM152 atlases. *Sci Data*. 2019;6(1):210. doi:10.1038/s41597-019-0217-0

23.	Nieuwenhuys R. *The Human Central Nervous System*. Accessed November 9, 2022. https://link.springer.com/book/10.1007/978-3-540-34686-9

24.	Hazrati LN, Parent A. The striatopallidal projection displays a high degree of anatomical specificity in the primate. *Brain Res*. 1992;592(1-2):213-227. doi:10.1016/0006-8993(92)91679-9

25.	Hedreen JC, Delong MR. Organization of striatopallidal, striatonigral, and nigrostriatal projections in the macaque. *Journal of Comparative Neurology*. 1991;304(4):569-595. doi:10.1002/cne.903040406

26.	Percheron G, Yelnik J, François C. A Golgi analysis of the primate globus pallidus. III. Spatial organization of the striato-pallidal complex. *J Comp Neurol*. 1984;227(2):214-227. doi:10.1002/cne.902270207

27.	François C, Yelnik J, Percheron G, Fénelon G. Topographic distribution of the axonal endings from the sensorimotor and associative striatum in the macaque pallidum and substantia nigra. *Exp Brain Res*. 1994;102(2):305-318. doi:10.1007/BF00227517

28.	Haber SN, Fudge JL, McFarland NR. Striatonigrostriatal Pathways in Primates Form an Ascending Spiral from the Shell to the Dorsolateral Striatum. *J Neurosci*. 2000;20(6):2369-2382. doi:10.1523/JNEUROSCI.20-06-02369.2000

29.	Lynd-Balta E, Haber SN. Primate striatonigral projections: A comparison of the sensorimotor-related striatum and the ventral striatum. *Journal of Comparative Neurology*. 1994;345(4):562-578. doi:10.1002/cne.903450407

30.	Haber SN, Groenewegen HJ, Grove EA, Nauta WJH. Efferent connections of the ventral pallidum: Evidence of a dual striato pallidofugal pathway. *Journal of Comparative Neurology*. 1985;235(3):322-335. doi:10.1002/cne.902350304

31.	Haber SN, Lynd E, Klein C, Groenewegen HJ. Topographic organization of the ventral striatal efferent projections in the rhesus monkey: An anterograde tracing study. *Journal of Comparative Neurology*. 1990;293(2):282-298. doi:10.1002/cne.902930210

32.	Spooren W p. j. m., Lynd-Balta E, Mitchell S, Haber S n. Ventral pallidostriatal pathway in the monkey: Evidence for modulation of basal ganglia circuits. *Journal of Comparative Neurology*. 1996;370(3):295-312. doi:10.1002/(SICI)1096-9861(19960701)370:3%3C295::AID-CNE2%3E3.0.CO;2-%23

33.	François C, Percheron G, Parent A, Sadikot AF, Fenelon G, Yelnik J. Topography of the projection from the central complex of the thalamus to the sensorimotor striatal territory in monkeys. *Journal of Comparative Neurology*. 1991;305(1):17-34. doi:10.1002/cne.903050104

34.	Ilyas A, Pizarro D, Romeo AK, Riley KO, Pati S. The centromedian nucleus: Anatomy, physiology, and clinical implications. *Journal of Clinical Neuroscience*. 2019;63:1-7. doi:https://doi.org/10.1016/j.jocn.2019.01.050

35.	Sadikot AF, Rymar VV. The primate centromedian–parafascicular complex: Anatomical organization with a note on neuromodulation. *Brain Research Bulletin*. 2009;78(2):122-130. doi:https://doi.org/10.1016/j.brainresbull.2008.09.016

36.	Sadikot AF, Parent A, Smith Y, Bolam JP. Efferent connections of the centromedian and parafascicular thalamic nuclei in the squirrel monkey: A light and electron microscopic study of the thalamostriatal projection in relation to striatal heterogeneity. *The Journal of Comparative Neurology*. 1992;320(2):228-242. doi:10.1002/cne.903200207

37.	Sadikot AF, Parent A, François C. Efferent connections of the centromedian and parafascicular thalamic nuclei in the squirrel monkey: A PHA-L study of subcortical projections. *Journal of Comparative Neurology*. 1992;315(2):137-159. doi:10.1002/cne.903150203

38.	Parent M, Parent A. Single-axon tracing and three-dimensional reconstruction of centre médian-parafascicular thalamic neurons in primates. *Journal of Comparative Neurology*. 2005;481(1):127-144. doi:10.1002/cne.20348

39.	Alho EJL, Alho ATDL, Horn A, et al. The Ansa Subthalamica: A Neglected Fiber Tract. *Movement Disorders*. 2020;35(1):75-80. doi:10.1002/mds.27901

40.	Li M, Ribas EC, Zhang Z, et al. Tractography of the ansa lenticularis in the human brain. *Clinical Anatomy*. 2022;35(3):269-279. doi:10.1002/ca.23788

41.	Chung BS, Park JS. Whole course of pallidothalamic tracts identified on the sectioned images and surface models. *Clin Anat*. 2020;33(1):66-76. doi:10.1002/ca.23468

42.	Hazrati LN, Parent A. Projection from the external pallidum to the reticular thalamic nucleus in the squirrel monkey. *Brain Res*. 1991;550(1):142-146. doi:10.1016/0006-8993(91)90418-u

43.	Kim R, Nakano K, Jayaraman A, Carpenter MB. Projections of the globus pallidus and adjacent structures: An autoradiographic study in the monkey. *Journal of Comparative Neurology*. 1976;169(3):263-289. doi:10.1002/cne.901690302

44.	Neudorfer C, Maarouf M. Neuroanatomical background and functional considerations for stereotactic interventions in the H fields of Forel. *Brain Struct Funct*. 2018;223(1):17-30. doi:10.1007/s00429-017-1570-4

45.	Parent M, Parent A. The pallidofugal motor fiber system in primates. *Parkinsonism & Related Disorders*. 2004;10(4):203-211. doi:10.1016/j.parkreldis.2004.02.007

46.	Baron MS, Sidibé M, DeLong MR, Smith Y. Course of motor and associative pallidothalamic projections in monkeys. *Journal of Comparative Neurology*. 2001;429(3):490-501. doi:10.1002/1096-9861(20010115)429:3%3C490::AID-CNE9%3E3.0.CO;2-K

47.	Parent M, Lévesque M, Parent A. Two types of projection neurons in the internal pallidum of primates: Single-axon tracing and three-dimensional reconstruction. *Journal of Comparative Neurology*. 2001;439(2):162-175. doi:10.1002/cne.1340

48.	Sakai ST, Inase M, Tanji J. Comparison of cerebellothalamic and pallidothalamic projections in the monkey (Macaca fuscata): A double anterograde labeling study. *Journal of Comparative Neurology*. 1996;368(2):215-228. doi:10.1002/(SICI)1096-9861(19960429)368:2%3C215::AID-CNE4%3E3.0.CO;2-6

49.	Mehler WR, Nauta WJ. Connections of the basal ganglia and of the cerebellum. *Confin Neurol*. 1974;36(4-6):205-222. doi:10.1159/000102797

50.	Mai J, Majtanik M, Paxinos G. *Atlas of the Human Brain (4th Edition)*. Academic Press; 2015.

51.	Middleton FA, Strick PL. Cerebellar Output Channels. In: Schmahmann JD, ed. *International Review of Neurobiology*. Vol 41. Review of. Academic Press; 1997:61-82. doi:10.1016/S0074-7742(08)60347-5

52.	Rouiller EM, Liang F, Babalian A, Moret V, Wiesendanger M. Cerebellothalamocortical and pallidothalamocortical projections to the primary and supplementary motor cortical areas: A multiple tracing study in macaque monkeys. *Journal of Comparative Neurology*. 1994;345(2):185-213. doi:10.1002/cne.903450204

53.	Middleton FA, Strick PL. Basal ganglia and cerebellar loops: motor and cognitive circuits. *Brain Research Reviews*. 2000;31(2):236-250. doi:10.1016/S0165-0173(99)00040-5

54.	Sakai ST, Stepniewska I, Qi HX, Kaas JH. Pallidal and cerebellar afferents to pre-supplementary motor area thalamocortical neurons in the owl monkey: a multiple labeling study. *J Comp Neurol*. 2000;417(2):164-180.

55.	Mason A, Ilinsky IA, Maldonado S, Kultas-Ilinsky K. Thalamic terminal fields of individual axons from the ventral part of the dentate nucleus of the cerebellum in Macaca mulatta. *Journal of Comparative Neurology*. 2000;421(3):412-428. doi:10.1002/(SICI)1096-9861(20000605)421:3%3C412::AID-CNE9%3E3.0.CO;2-Z

56.	Stanton GB. Topographical organization of ascending cerebellar projections from the dentate and interposed nuclei in Macaca mulatta: An anterograde degeneration study. *Journal of Comparative Neurology*. 1980;190(4):699-731. doi:10.1002/cne.901900406

57.	Hintzen A, Pelzer EA, Tittgemeyer M. Thalamic interactions of cerebellum and basal ganglia. *Brain Struct Funct*. 2018;223(2):569-587. doi:10.1007/s00429-017-1584-y

58.	Berkley KJ. Spatial relationships between the terminations of somatic sensory motor pathways in the rostral brainstem of cats and monkeys. II. Cerebellar projections compared with those of the ascending somatic sensory pathways in lateral diencephalon. *Journal of Comparative Neurology*. 1983;220(2):229-251. doi:10.1002/cne.902200210

59.	Berkley KJ. Spatial relationships between the terminations of somatic sensory and motor pathways in the rostral brainstem of cats and monkeys. I. Ascending somatic sensory inputs to lateral diencephalon. *Journal of Comparative Neurology*. 1980;193(1):283-317. doi:10.1002/cne.901930119

60.	Kaas JH, Nelson RJ, Sur M, Dykes RW, Merzenich MM. The somatotopic organization of the ventroposterior thalamus of the squirrel monkey, Saimiri sciureus. *Journal of Comparative Neurology*. 1984;226(1):111-140. doi:10.1002/cne.902260109

61.	Lemon RN, Morecraft RJ. The evidence against somatotopic organization of function in the primate corticospinal tract. *Brain*. 2023;146(5):1791-1803. doi:10.1093/brain/awac496

62.	Meola A, Yeh FC, Fellows-Mayle W, Weed J, Fernandez-Miranda JC. Human Connectome-Based Tractographic Atlas of the Brainstem Connections and Surgical Approaches. *Neurosurgery*. 2016;79(3):437-455. doi:10.1227/NEU.0000000000001224

63.	Pauli WM, Nili AN, Tyszka JM. A high-resolution probabilistic in vivo atlas of human subcortical brain nuclei. *Sci Data*. 2018;5(1):180063. doi:10.1038/sdata.2018.63

64.	Ewert S, Plettig P, Li N, et al. Toward defining deep brain stimulation targets in MNI space: A subcortical atlas based on multimodal MRI, histology and structural connectivity. *NeuroImage*. 2018;170:271-282. doi:10.1016/j.neuroimage.2017.05.015

65.	Neudorfer C, Germann J, Elias GJB, Gramer R, Boutet A, Lozano AM. A high-resolution in vivo magnetic resonance imaging atlas of the human hypothalamic region. *Sci Data*. 2020;7(1):305. doi:10.1038/s41597-020-00644-6

66.	Pijnenburg R, Scholtens LH, Ardesch DJ, de Lange SC, Wei Y, van den Heuvel MP. Myelo- and cytoarchitectonic microstructural and functional human cortical atlases reconstructed in common MRI space. *NeuroImage*. 2021;239:118274. doi:10.1016/j.neuroimage.2021.118274

67.	Haber SN, Calzavara R. The cortico-basal ganglia integrative network: the role of the thalamus. *Brain Res Bull*. 2009;78(2-3):69-74. doi:10.1016/j.brainresbull.2008.09.013
